# Feasibility and Psychological Outcomes of a Clinical Meditation Intervention for Patients with Cancer: A Mixed-Methods Pilot Study

**DOI:** 10.64898/2026.11.11.26367801

**Authors:** Atsushi Niida, Tomoko Koshiyama, Keisuke Baba, Gota Fujisawa, Naomi Iwane, Daien Ohshita, Dai Shida, Narikazu Boku

**Affiliations:** Laboratory of Molecular Medicine, Human Genome Center, The Institute of Medical Science, The University of Tokyo, Tokyo, Japan; Clinical Meditation Education and Research Institute, Hida-Senkoji Temple, Takayama, Japan; Department of Oncology and General Medicine, IMSUT Hospital, The Institute of Medical Science, The University of Tokyo, Tokyo, Japan; School of Health and Nursing Science, Wakayama Medical University, Wakayama, Japan; Division of Frontier Surgery, The Institute of Medical Science, The University of Tokyo, Tokyo, Japan

**Keywords:** cancer patients, meditation, spiritual well-being, posttraumatic growth, mindfulness, qualitative research, thematic analysis, convergent mixed-methods design

## Abstract

**Background:** Psychological care is an important component of cancer care, addressing psychological distress and supporting patients as they cope with illness. Meditation-based approaches, including mindfulness-based interventions (MBIs), have been incorporated into supportive cancer care internationally. Clinical Meditation (Rinsho Meiso-ho), a meditation method developed in Japan, integrates traditional Buddhist meditation with a contemporary medical care perspective. However, its feasibility and psychological outcomes in patients with cancer have not been sufficiently studied.

**Objective:** This study aimed to evaluate the feasibility of a Clinical Meditation intervention and to explore psychological outcomes and participants’ experiences among patients with cancer who were receiving or scheduled to receive pharmacotherapy at the time of recruitment.

**Methods:** This single-arm pilot study used a convergent mixed-methods design to evaluate an 8-week program consisting of a 4-week structured phase (1 face-to-face session and 3 brief online group sessions, with prescribed home practice) and a 4-week self-directed phase (1 individual online session, with voluntary home practice). Feasibility was assessed descriptively using retention, attendance, practice frequency, and poststudy feedback. Anxiety, depression, insomnia, fatigue, spiritual well-being (SWB), and posttraumatic growth (PTG) were assessed at baseline, 4 weeks, and 8 weeks. Changes were examined using Friedman tests and exact Wilcoxon signed-rank tests, with adjustment for multiple comparisons. Free-text responses were analyzed thematically and integrated with questionnaire findings at the participant level.

**Results:** Eleven patients were enrolled; 9 (81.8%) completed the 4-week assessment and 8 (72.7%) completed the 8-week assessment. Median practice frequency declined from 5–6 days per week during the structured first month to 3–4 days per week during the self-directed second month (P=.031). Quantitative analyses included 9 participants, of whom 8 completed all 3 assessments; qualitative analyses included 8 participants. SWB and PTG showed significant overall changes (adjusted P=.02 and P=.04, respectively). SWB was higher than baseline at 4 weeks (adjusted P=.012), but the difference was not significant at 8 weeks. PTG was higher than baseline at both 4 and 8 weeks (both adjusted P=.047). No significant changes were observed in anxiety, depression, insomnia, or fatigue. Five themes were identified, encompassing experiences such as relaxation, greater distance from rumination, and meditation as a source of support. Participants’ accounts generally aligned with increases in SWB and PTG scores, although perceived benefits did not always correspond to changes in all questionnaire measures.

**Conclusions:** This mixed-methods pilot study suggests that Clinical Meditation may be feasible and acceptable for patients with cancer, although sustaining meditation practice remains challenging. Exploratory increases in SWB and PTG warrant further investigation. Larger controlled studies, comparisons with established MBIs, and deeper qualitative research are needed to evaluate outcomes and clarify the distinctive features of Clinical Meditation.

## Introduction

During diagnosis and treatment, patients with cancer frequently experience psychological distress, including anxiety and depression, as well as related symptoms such as insomnia and fatigue, which can adversely affect quality of life (QOL). Psycho-oncologic interventions have been associated with small-to-moderate improvements in emotional distress and QOL [1,2].

Among these, meditation-based approaches, particularly mindfulness-based interventions (MBIs) such as mindfulness-based stress reduction (MBSR) and mindfulness-based cognitive therapy (MBCT), have been among the most extensively studied complementary approaches to symptom management in patients with cancer. Meta-analyses of randomized controlled trials have shown that MBIs reduce anxiety and depression [3,4] and alleviate cancer-related fatigue [5]. For sleep, MBIs have been associated with improved sleep quality, although evidence for reducing insomnia severity remains limited [4,6]. On the basis of this evidence, the joint guideline from the Society for Integrative Oncology and the American Society of Clinical Oncology strongly recommends MBIs for reducing anxiety and depressive symptoms in patients with cancer [1].

Beyond symptom relief, meditation may also support positive dimensions of psychological adjustment. A cancer diagnosis can challenge patients’ sense of meaning and raise existential questions about life and death. Spiritual well-being (SWB) encompasses a sense of meaning, peace, and, for some patients, faith, and has been associated with better QOL in patients with cancer [7]. Posttraumatic growth (PTG) refers to positive psychological changes that may emerge through the struggle with a major life crisis, including a greater appreciation of life, closer relationships, new possibilities, personal strength, and spiritual change [8]. Such growth can coexist with psychological distress and is not simply the absence of symptoms. A meta-analysis of randomized controlled trials of MBIs in patients with cancer reported improvements in spiritual outcomes and personal growth [9]. A more recent meta-analysis focusing on perceived PTG also found a moderate pooled effect of mindfulness- and acceptance-based interventions in cancer populations [10]. However, the limited number of trials, their heterogeneity, and risk of bias warrant cautious interpretation. These findings support examining SWB and PTG as exploratory outcomes of meditation interventions that address existential concerns and the meaning of illness.

In Japan, an 8-week randomized controlled trial of MBCT in patients with breast cancer reported improvements in psychological distress, SWB, and QOL compared with a wait-list control group [11]. A small uncontrolled study reported reduced psychological distress following a brief mindfulness-based meditation program in Japanese patients receiving cancer treatment [12].

Nevertheless, evidence on mindfulness and meditation in Japanese cancer populations remains limited. A 2020 review also noted that MBIs had not been widely integrated into oncology practice in Japan [13]. Moreover, the time demands of participation may hinder clinical implementation. Standard MBSR programs typically involve weekly sessions over approximately 8 weeks and regular home practice [14,15]. In a study of MBSR for women with metastatic breast cancer, the 8-week time commitment was cited as a reason for nonparticipation and was challenging for those who participated [15]. These considerations highlight the need to evaluate shorter, less demanding meditation interventions that are feasible, acceptable, and manageable alongside cancer treatment in clinical settings.

Clinical Meditation (Rinsho Meiso-ho), a meditation method developed in Japan by Daien Ohshita, integrates traditional Buddhist meditation with a contemporary medical care perspective [16] (see Multimedia Appendix 1 for a detailed description of the method and its background). It can be practiced regardless of religious affiliation or belief and incorporates elements that engage directly with spiritual questions concerning life, death, and the meaning of life. By offering opportunities to explore these questions and reflect on the experience of illness, Clinical Meditation may support SWB and aspects of PTG, such as appreciation of life and spiritual change, among patients with cancer. A simplified meditation guidance program based on this method was previously developed, and its preliminary feasibility was reported in a small pilot study of patients with mild cognitive impairment [17]. However, evidence in patients with cancer remains limited, and its psychological effects, including those on SWB and PTG, warrant further investigation.

Mixed-methods designs, which combine questionnaire outcomes with participants’ accounts, allow both psychological changes and patients’ experiences of an intervention to be examined. Previous mixed-methods studies of MBIs in patients with cancer have shown that qualitative interviews can reveal perceived benefits, such as increased awareness, acceptance, and personal growth, that may not be fully captured by questionnaire outcomes [18,19]. A convergent mixed-methods design extends this approach by analyzing quantitative and qualitative data separately and then integrating the findings to examine whether they confirm, expand, or are discordant with each other [20,21]. Using this design, Eyles et al examined the feasibility and acceptability of an 8-week MBSR program for women with metastatic breast cancer by combining questionnaire assessments with qualitative accounts from patients and health care professionals [15].

Against this background, we conducted a single-arm pilot study using a convergent mixed-methods design among patients with cancer who were receiving or scheduled to receive pharmacotherapy. To reduce the burden of participation, the intervention concentrated structured guidance into the first 4 weeks—an initial face-to-face session followed by 3 brief weekly online group sessions—and was followed by a self-directed month that included 1 individual online session. The primary objective was to evaluate the feasibility of a Clinical Meditation guidance intervention; the secondary, exploratory objectives were to examine psychological outcomes and participants’ experiences.

## Methods

### Participants and Recruitment

Eligible participants were patients with cancer aged 20 years or older who were receiving outpatient care at IMSUT Hospital, The Institute of Medical Science, The University of Tokyo, and who were receiving or scheduled to receive pharmacotherapy. Patients with severe psychiatric illness were excluded. Participants were recruited on a rolling basis through flyers distributed by attending physicians and posted within the hospital, each containing a QR code linking to the study explanation document. In addition, the principal investigator provided direct explanations of the study as needed. The planned recruitment target was 10 participants; as this was a pilot study, the sample size was not based on a formal power calculation.

### Ethical Considerations

This study was approved by the Institutional Review Board of the Institute of Medical Science, The University of Tokyo (approval number: 2025-3-0501). Online informed consent was obtained after participants had reviewed and understood the study explanation document. Participants were free to withdraw at any time. Participants received no financial or other compensation for their participation. Participant names were replaced with coded identifiers (eg, Patient_A) before analysis, and the key linking these identifiers to participants’ identities was stored separately from the study data.

### Intervention Program

The intervention comprised a 4-week structured guidance phase followed by a 4-week self-directed phase (Figure 1) and included 3 types of sessions: (1) an initial face-to-face group session (approximately 1 hour) in a seminar room at the Institute of Medical Science, The University of Tokyo, including a lecture on Clinical Meditation and practical instruction centered on “Loosening Meditation” (Yurumeru Meiso); (2) 3 weekly online group sessions (approximately 30 minutes each) involving meditation practice and group sharing; and (3) an individual online session (approximately 30 minutes) conducted 5–7 weeks after the initial session, providing individualized guidance and an opportunity to discuss personal concerns.

**Figure 1.**
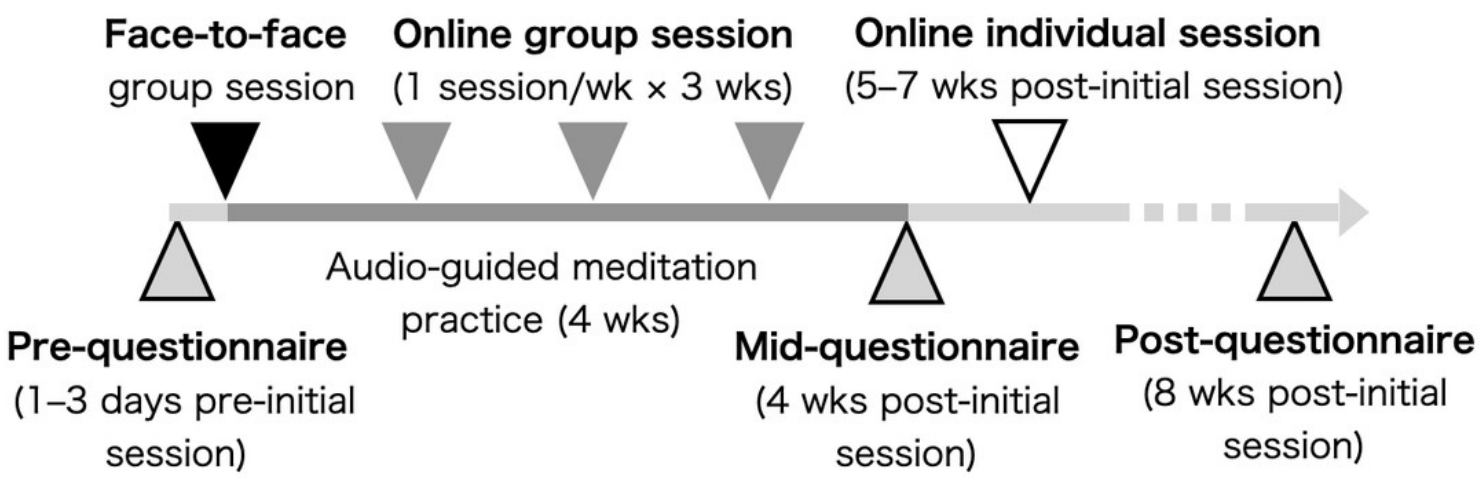
Timeline of the intervention and questionnaire assessments. Patients with cancer who were receiving or scheduled to receive pharmacotherapy at recruitment participated in an 8-week Clinical Meditation guidance intervention. The intervention comprised a 4-week structured phase (1 face-to-face session and 3 brief online group sessions, with prescribed home practice) followed by a 4-week self-directed phase (1 individual online session, with voluntary home practice). Outcomes were assessed at 3 time points: before the intervention (Pre), at 4 weeks (Mid), and at 8 weeks (Post).

The program was designed by Tomoko Koshiyama (TK) and Daien Ohshita (DO). Meditation guidance was delivered by TK, an experienced Clinical Meditation instructor, under the supervision of DO. As participants enrolled, small groups of 1–3 were formed for the initial face-to-face session and subsequent online group sessions. The principal investigator, Atsushi Niida (AN), attended the initial and online group sessions, but not the individual sessions.

During the first month, participants were asked to practice meditation for 15 minutes, using a YouTube audio guide, on days without scheduled sessions. During the second month, continued practice was encouraged but left to participants’ discretion. Further details of the Clinical Meditation method, theoretical background, and session protocols are provided in Multimedia Appendix 1.

### Questionnaire Data Collection

Participants completed a web-based questionnaire at 3 time points (Figure 1): 1–3 days before the initial session (Pre), 4 weeks after the initial session (28–31 days; Mid), and 8 weeks after the initial session (54–57 days; Post). Six psychological outcomes—anxiety, depression, insomnia, fatigue, spiritual well-being (SWB), and posttraumatic growth (PTG)—were assessed using the Japanese versions of the following 5 questionnaires.

- Anxiety and depression: the Hospital Anxiety and Depression Scale (HADS), comprising a 7-item anxiety subscale (HADS-A) and a 7-item depression subscale (HADS-D), each scored from 0 to 21. Conventional cutoffs classify subscale scores of 0–7 as noncases, 8–10 as borderline cases, and 11–21 as probable cases [22]. The Japanese version has been evaluated in patients with cancer [23].
- Insomnia: the Insomnia Severity Index (ISI), a 7-item questionnaire assessing insomnia severity (0–28 points). Scores of 0–7 indicate no clinically significant insomnia, 8–14 subthreshold insomnia, 15–21 moderate insomnia, and 22–28 severe insomnia [24,25].
- Fatigue: the Brief Fatigue Inventory (BFI), comprising 9 items assessing fatigue severity and its interference with daily functioning. Each item is scored from 0 to 10, and the global score is calculated as the mean of the completed items, provided that at least 5 of the 9 items are answered, per the original scoring rule [26,27].
- SWB: the 12-item Functional Assessment of Chronic Illness Therapy-Spiritual (FACIT-Sp-12), with scores ranging from 0 to 48 and higher scores indicating better SWB [7,28].
- PTG: the Japanese version of the Short Form of the Expanded Version of the Posttraumatic Growth Inventory (PTGI-X-SF-J), comprising 10 items scored from 0 to 5 [29]. In this study, the mean item score (0–5) was used.

In addition, at the Mid and Post assessments, participants rated their frequency of meditation practice during the preceding month using a 5-point categorical scale (0, 1–2, 3–4, 5–6, or 7 days per week), with attendance at the meditation sessions counted as meditation practice.

After completion of the Post assessment, participants were invited to complete an additional, anonymous online questionnaire assessing their experience of study participation, administered separately from the Pre/Mid/Post outcome measures described above and not linked to individual participant identifiers. This questionnaire asked participants to rate, on a 0–10 scale, their satisfaction with the face-to-face session, the online group sessions, and the individual online session; the burden associated with completing the questionnaires and with daily home meditation practice (higher scores indicating less burden); their satisfaction with the administrative support provided during the study; their willingness to continue meditation practice; and their overall satisfaction with study participation. Because responses were collected anonymously, they could not be linked to individual participants or to their Pre/Mid/Post questionnaire data.

### Feasibility Evaluation

Feasibility was evaluated descriptively using retention at the Mid and Post assessments, attendance at each scheduled session, and self-reported meditation practice frequency. Retention and attendance were summarized as counts and percentages using all enrolled participants as the denominator, with additional summaries restricted to those who continued beyond the initial session. Acceptability was assessed using the anonymous poststudy questionnaire described above. No criteria for determining feasibility were prespecified.

### Statistical Analysis of Questionnaire Data

For each of the 6 outcome scores derived from the 5 questionnaires, overall change across the 3 time points (Pre, Mid, and Post) was assessed using the Friedman test, including only participants with complete data at all 3 time points. No single primary outcome was prespecified; all 6 measures were assessed on an equal, exploratory basis. To account for testing 6 outcome measures, Friedman P values were adjusted for multiplicity using the Benjamini-Hochberg procedure [30] (adjusted P<.05). For outcome measures with a significant Friedman result, post hoc pairwise comparisons between time points were conducted using the 2-tailed Wilcoxon signed-rank test using the exact conditional sign-flip distribution of the observed absolute paired differences; because these 3 comparisons (Pre-Mid, Mid-Post, Pre-Post) constitute all pairwise comparisons among the 3 time points, the resulting P values were adjusted for multiplicity within each outcome using Shaffer’s modified sequentially rejective procedure [31]. Each pairwise comparison included participants with data at both corresponding time points. Zero paired differences were excluded from the signed-rank statistic, and tied absolute differences were assigned average ranks. Paired differences were rounded to 10 decimal places before ranking to avoid numerical artifacts in the handling of ties. As the effect-size measure for the pairwise comparisons, we calculated the matched-pairs rank-biserial correlation (r), which is derived directly from the Wilcoxon signed-rank statistic [32]. The Hodges-Lehmann estimate was calculated as the median of the Walsh averages of all paired differences, including zero differences. Confidence intervals were obtained by inverting the same 2-tailed exact signed-rank test across hypothesized location shifts; differences and ranks were recalculated at each shift, and the closure of the nonrejected set was reported. A location-shift interpretation assumes symmetry of the paired-difference distribution. Full Friedman statistics for all 6 outcome measures, together with medians, interquartile ranges, Hodges-Lehmann estimates of the paired location shift (the pseudomedian of paired differences) with unadjusted 95% CIs, and post hoc exact Wilcoxon results, are provided in Tables S1 and S2 in Multimedia Appendix 2.

To explore how the outcome measures co-varied, we estimated correlations among the 6 measures at 2 levels. Within-person associations were estimated using repeated measures correlation (rmcorr) [33], which removes between-participant differences in overall score levels; this analysis used all available observations. Between-person associations were estimated using Pearson correlation of each participant’s mean score across time points. Correlation matrices ordered by hierarchical clustering, correlation coefficients with 95% CIs obtained using the Fisher z transformation, and scatter plots are provided in Figure S4 in Multimedia Appendix 2. Statistical analyses were performed in R (version 4.5). All analysis code was generated with the assistance of large language models (LLMs; Claude Opus 4.8, 5.0, and 5.5; Anthropic) and was executed, reviewed, and verified by AN.

### Qualitative Analysis and Mixed-Methods Integration

In addition to the questionnaire items, participants provided free-text responses to 2 open-ended questions regarding their psychological state and any insights gained through meditation practice. Q1 (“In addition to the above questions, please freely describe anything you have noticed recently about your own state of mind.”) was administered at all 3 time points (Pre, Mid, and Post), whereas Q2 (“Please freely describe any insights you have gained, or changes you have felt, through your meditation practice.”) was administered only at the latter 2 time points (Mid and Post).

Thematic analysis followed Braun and Clarke’s 6-phase framework [34] and was conducted with assistance from an LLM. Before analysis, personally identifying information was manually removed from the Japanese-language free-text responses. The LLM (Claude Opus 4.8, Anthropic), configured to prevent the submitted data from being used for model training, was instructed to identify meaningful text units; generate candidate codes; compile a codebook containing code definitions, inclusion and exclusion criteria, and all text passages assigned to each code; and organize the codes into candidate themes based on their semantic relationships without weighting by code frequency.

AN then reviewed the LLM-generated codes, candidate themes, and supporting quotations against the original free-text responses, revised and consolidated codes, reconsidered themes as needed, and made the final decisions regarding the codebook and thematic structure. The finalized codebook and themes were reviewed by Naomi Iwane (NI), a qualitative research methodologist who was not involved in the initial coding. Her feedback informed further refinements but did not constitute independent recoding or a comprehensive audit.

Regarding researcher reflexivity, both AN and NI had completed the Clinical Meditation instructor training program, providing them with theoretical and practical familiarity with Clinical Meditation that informed their interpretation but may also have predisposed them toward interpretations favorable to the method. In addition, AN was involved in participant recruitment and attended the group sessions (see Intervention Program). All stages of thematic analysis were conducted in Japanese. The resulting themes and coded passages were subsequently translated into English with LLM assistance, and all translations were checked by AN. Reporting was guided by the Standards for Reporting Qualitative Research (SRQR) [35] (Multimedia Appendix 3). The final codebook, including the thematic structure and English translations of all coded passages, is provided in Multimedia Appendix 4.

Consistent with the convergent mixed-methods design, questionnaire data and qualitative free-text responses were collected at the same time points and analyzed separately [20]. The findings were then integrated at the individual participant level in a joint display to explore correspondence and divergence between participants’ reported experiences and changes in questionnaire scores [21]. Contextual information that participants shared during the group sessions, observed by AN, was used only to aid the interpretation of individual cases.

## Results

### Participant Characteristics and Feasibility

We enrolled 11 participants, whose cancer types included colorectal cancer (n=7; colon, n=5; rectum, n=2), gastric cancer (n=2), endometrial cancer (n=1), and duodenal cancer (n=1); disease stage ranged from III to IV (Table 1). All participants except 1 (Patient_D), who had completed a course of adjuvant chemotherapy shortly before the intervention began, were undergoing pharmacotherapy at the start of the intervention. Four participants (Patient_D, Patient_H, Patient_I, and Patient_K) reported limited prior experience with meditation, such as meditation during yoga sessions or school classes, use of meditation apps, or brief participation in another meditation program; none reported an established, ongoing meditation practice (Table 1).

**Table 1.** Participant clinical characteristics (N=11).

| **ID** | **Sex** | **Age** | **Cancer type** | **Stage** | **Clinical / treatment notes** | **Meditation experience** | **Withdrawal/Data missing** |
|---|---|---|---|---|---|---|---|
| Patient_A | M | Early 40s | Rectal | III |  |  |  |
| Patient_B | M | Early 70s | Gastric | IV | Unstable clinical condition during the intervention period |  |  |
| Patient_C | F | Early 70s | Rectal | III |  |  |  |
| Patient_D | F | Late 30s | Colon | III | Chemotherapy completed just before Pre | Meditation during yoga sessions and use of meditation apps |  |
| Patient_E | F | Early 50s | Colon | IV |  |  |  |
| Patient_F | F | Late 50s | Endometrial | IV |  |  |  |
| Patient_G | F | Late 60s | Duodenal | IV |  |  | Discontinued after Mid |
| Patient_H | M | Late 40s | Colon | III |  | Use of meditation apps | No free-text responses |
| Patient_I | M | Late 30s | Colon | III |  | Participation in another meditation program; ~1 month of practice |  |
| Patient_J | M | Early 60s | Gastric | IV | Unstable clinical condition during the intervention period |  | Discontinued after the face-to-face session |
| Patient_K | F | Late 50s | Colon | IV |  | Meditation during school classes | Discontinued after the face-to-face session |

M: male; F: female; app: application software

All 11 participants completed the baseline (Pre) questionnaire and the initial face-to-face group session. Two participants discontinued after the initial session. One of them had an unstable clinical condition; however, the reasons for discontinuation were not confirmed for either participant. No additional data were collected from these 2 participants (Figure 2a). One additional participant discontinued after the Mid assessment for an unconfirmed reason. Among the 11 enrolled participants, 81.8% completed the Mid assessment and 72.7% completed the Post assessment; among the 9 participants who continued beyond the initial session, this corresponds to 100% completion at Mid and 88.9% completion at Post. Attendance among all 11 enrolled participants was 45% (5/11) for the first online group session, 64% (7/11) for the second, 73% (8/11) for the third, and 64% (7/11) for the individual online session; among the 9 continuing participants, these correspond to 55.6% (5/9), 77.8% (7/9), 88.9% (8/9), and 77.8% (7/9), respectively (Figure S1 in Multimedia Appendix 2).

**Figure 2.**
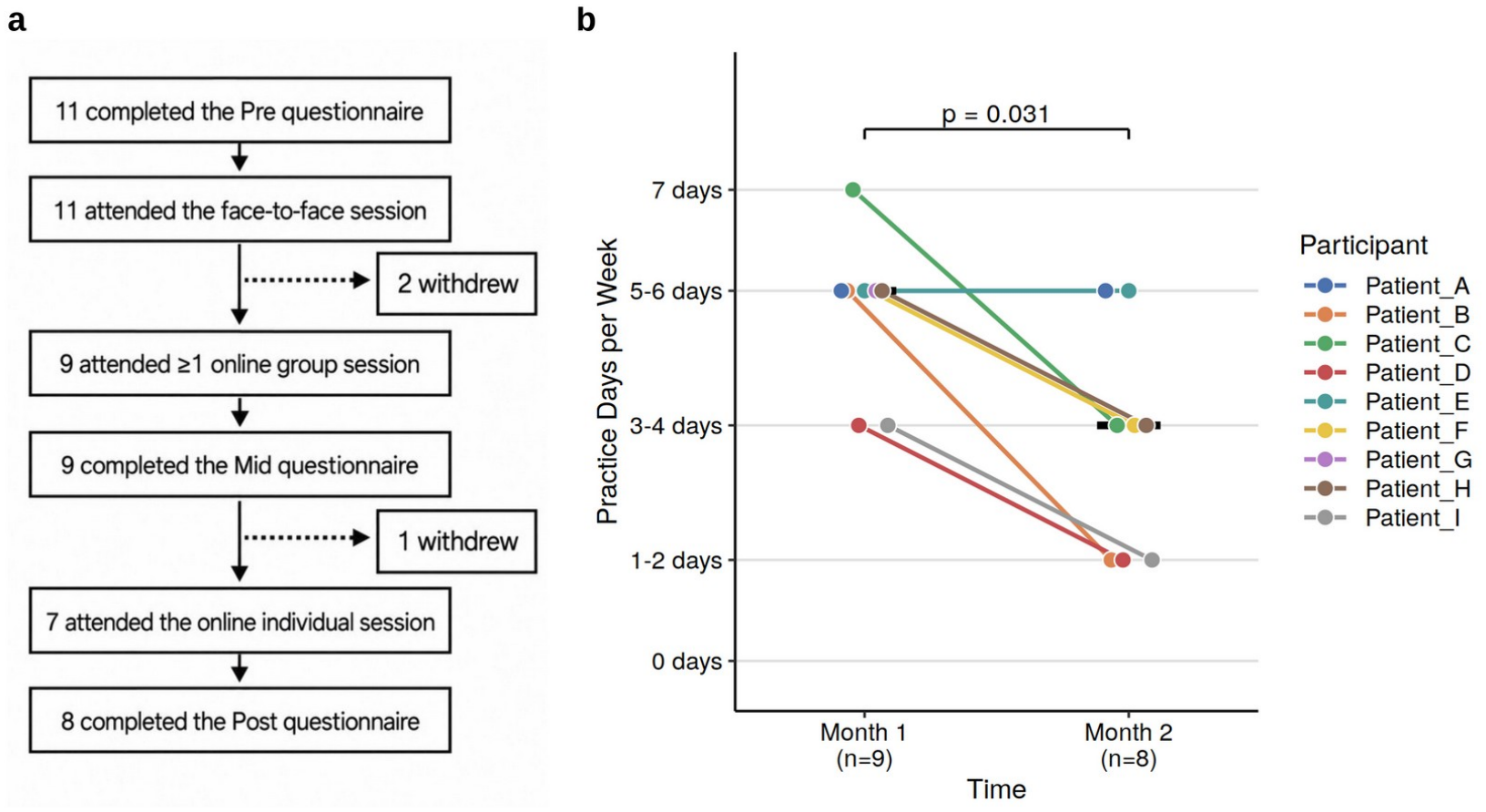
(a) Participant flow through the study. Session attendance and questionnaire completion were tracked independently; the 8 participants who completed the Post questionnaire did not necessarily attend every online session. (b) Meditation practice frequency by month. Frequency of meditation practice (days per week), including guided sessions and home practice, for each of the 9 participants at Month 1 and Month 2; Patient_G did not provide Month 2 data. Colored points connected by a line represent the same participant; black marks indicate the median. The *P* value was calculated using the exact Wilcoxon signed-rank test for the paired comparison (n=8).

The self-reported frequency of meditation practice, including both session attendance and home practice, was a median of 5–6 days per week during the first month of the intervention but significantly decreased to 3–4 days per week during the second month (Figure 2b; *P*=.031). This decline coincided with the shift from the first month, which included regular online sessions and prescribed audio-guided practice, to the second month, during which continuation of practice was left to participants’ own initiative.

Participants’ satisfaction with the intervention was assessed via an anonymous poststudy questionnaire completed by 6 of the 8 participants who completed the Post assessment (summarized in Figure S2 in Multimedia Appendix 2). On a 0–10 scale, the median score was 8 or higher for all assessed items, suggesting generally favorable participant experiences. Because responses were both anonymous and voluntary, response bias cannot be excluded, and these results should therefore be interpreted with caution.

Collectively, these findings provide preliminary information on participation and acceptability among patients who continued beyond the initial face-to-face session. However, attrition, declining practice frequency, and the absence of prespecified feasibility criteria limit conclusions about feasibility for the full enrolled cohort.

### Questionnaire Results

Statistical analyses of questionnaire data were conducted among the 9 participants who continued beyond the initial face-to-face session, excluding the 2 who discontinued at that point (Figure 3 and Figure S3 in Multimedia Appendix 2). Overall changes across the 3 time points were assessed for each outcome using the Friedman test in the 8 participants with complete data (Methods). Significant overall changes were observed for SWB (χ^2^=11.81, adjusted *P*=.02) and PTG (χ^2^=8.86, adjusted *P*=.04), whereas anxiety (HADS-A; χ^2^=5.43, adjusted *P*=.13), depression (HADS-D; χ^2^=0.48, adjusted *P*=.91), insomnia (ISI; χ^2^=3.27, adjusted *P*=.29), and fatigue (BFI; χ^2^=0.19, adjusted *P*=.91) did not show significant changes.

**Figure 3.**
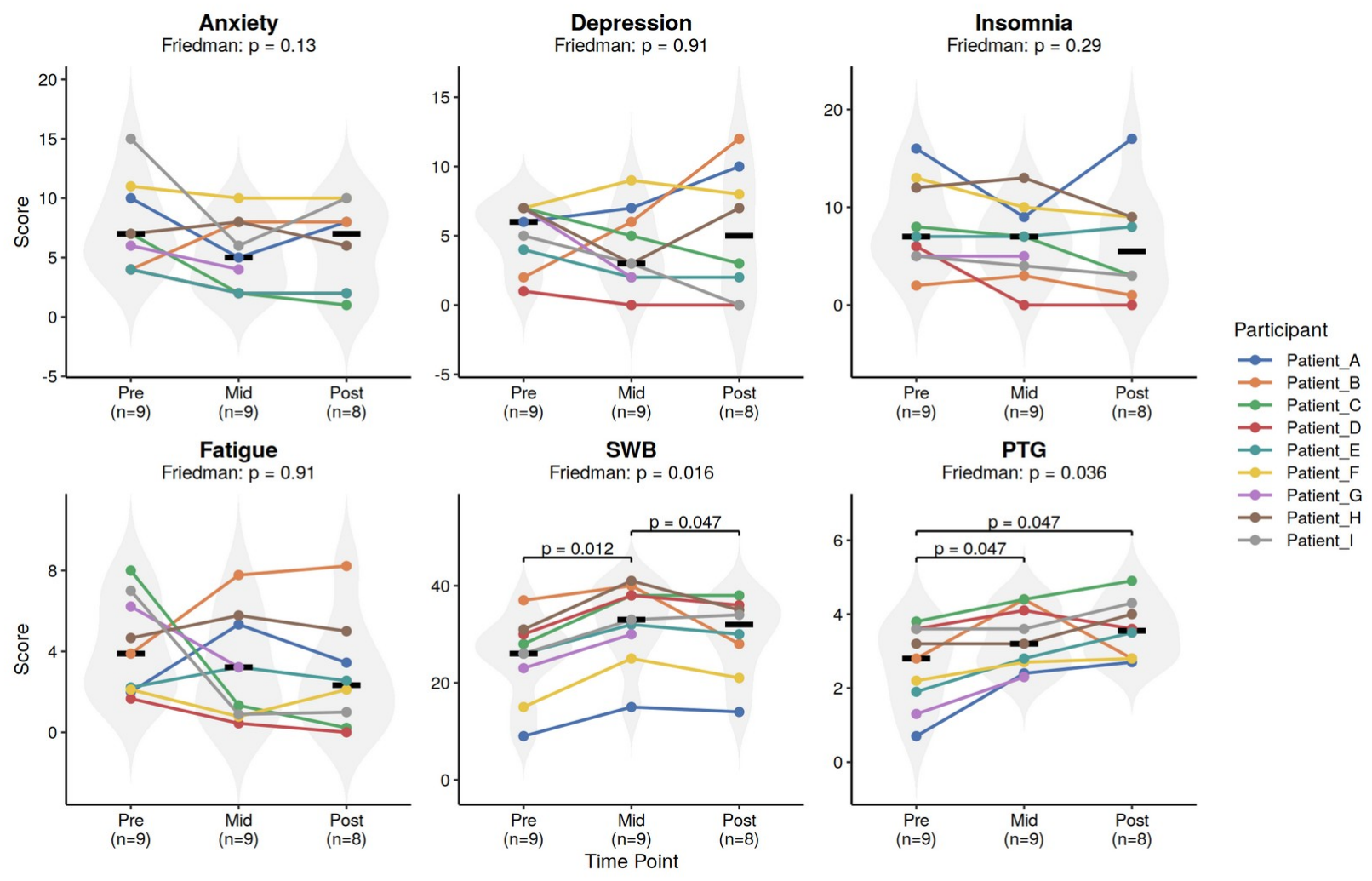
Changes in questionnaire scores over time. Violin plots show scores for 6 outcome measures at baseline (Pre), 4 weeks (Mid), and 8 weeks (Post): anxiety (HADS-A), depression (HADS-D), insomnia (ISI), fatigue (BFI), SWB (FACIT-Sp), and PTG (PTGI-X-SF-J) (n=9 at Pre and Mid; n=8 at Post). Colored points connected by a line represent the same participant, and black marks indicate the median at each time point. Each panel title reports the Friedman *P* value, adjusted for the 6 outcome measures using the Benjamini-Hochberg procedure, for the omnibus test of overall change across time points in the 8 participants with complete data. For SWB and PTG, the only 2 outcomes with a significant omnibus result, brackets report the Shaffer-adjusted *P* value from post hoc exact Wilcoxon signed-rank pairwise comparisons within that outcome.

For SWB and PTG, post hoc pairwise comparisons were performed using the exact Wilcoxon signed-rank test (Methods). SWB increased significantly from Pre to Mid (adjusted P=.012, r=1.00) and subsequently decreased from Mid to Post (adjusted P=.047, r=−0.89); the overall Pre-to-Post change was not significant (adjusted P=.125, r=0.61). PTG also increased significantly from Pre to Mid (adjusted P=.047, r=1.00) and remained significantly higher at Post than at Pre (adjusted P=.047, r=1.00), whereas the change from Mid to Post was not significant (adjusted P=.398, r=0.36).

Anxiety and insomnia scores showed decreasing trends from Pre to Post, whereas depression and fatigue showed no consistent direction of change, with similar numbers of participants showing decreases and increases; none of these 4 measures showed a significant overall change in the Friedman tests (full statistics are provided in Tables S1 and S2 in Multimedia Appendix 2). In a separate exploratory analysis, correlations among the 6 outcome measures (Figure S4 in Multimedia Appendix 2) showed that SWB and PTG were positively correlated at both the within-person (r=0.62, 95% CI 0.21-0.84) and between-person (r=0.77, 95% CI 0.22-0.95) levels, the only pair of the 15 examined whose 95% CIs excluded zero at both levels. In hierarchical clustering, SWB and PTG formed a cluster separate from the 4 symptom measures at both levels, and correlations between the 2 clusters were predominantly negative. Given the small sample and the number of pairs examined, these associations should be regarded as exploratory.

Overall, in this small sample, SWB and PTG were the only outcomes that changed significantly over time, and they were also positively correlated with each other, whereas the 4 symptom measures showed no significant change. Descriptively, however, their trajectories differed: the gain in SWB at Mid was not maintained at Post, whereas PTG remained above baseline.

### Qualitative Findings

Of the 9 participants who provided questionnaire data, 8 provided free-text responses at 1 or more time points. The participant who did not provide free-text responses (Patient_H) was not a native speaker of Japanese; although the reason for non-response was not confirmed, writing free-text responses in Japanese may have been difficult for this participant. In total, these 8 participants contributed 34 free-text responses across the Pre, Mid, and Post time points combined. Response length varied considerably across participants, ranging from 16 to 322 characters per response (mean 83 characters; Figure S5 in Multimedia Appendix 2). From these responses, 72 coded passages were extracted (Figure S6 in Multimedia Appendix 2). Qualitative analysis organized these participants’ narratives into 5 themes, described below; a subtheme was also identified within each of 2 themes (Theme 1 and Theme 3) and is described alongside its parent theme. The distribution of these themes across participants and time points is summarized visually in Figure 4a.

**Figure 4.**
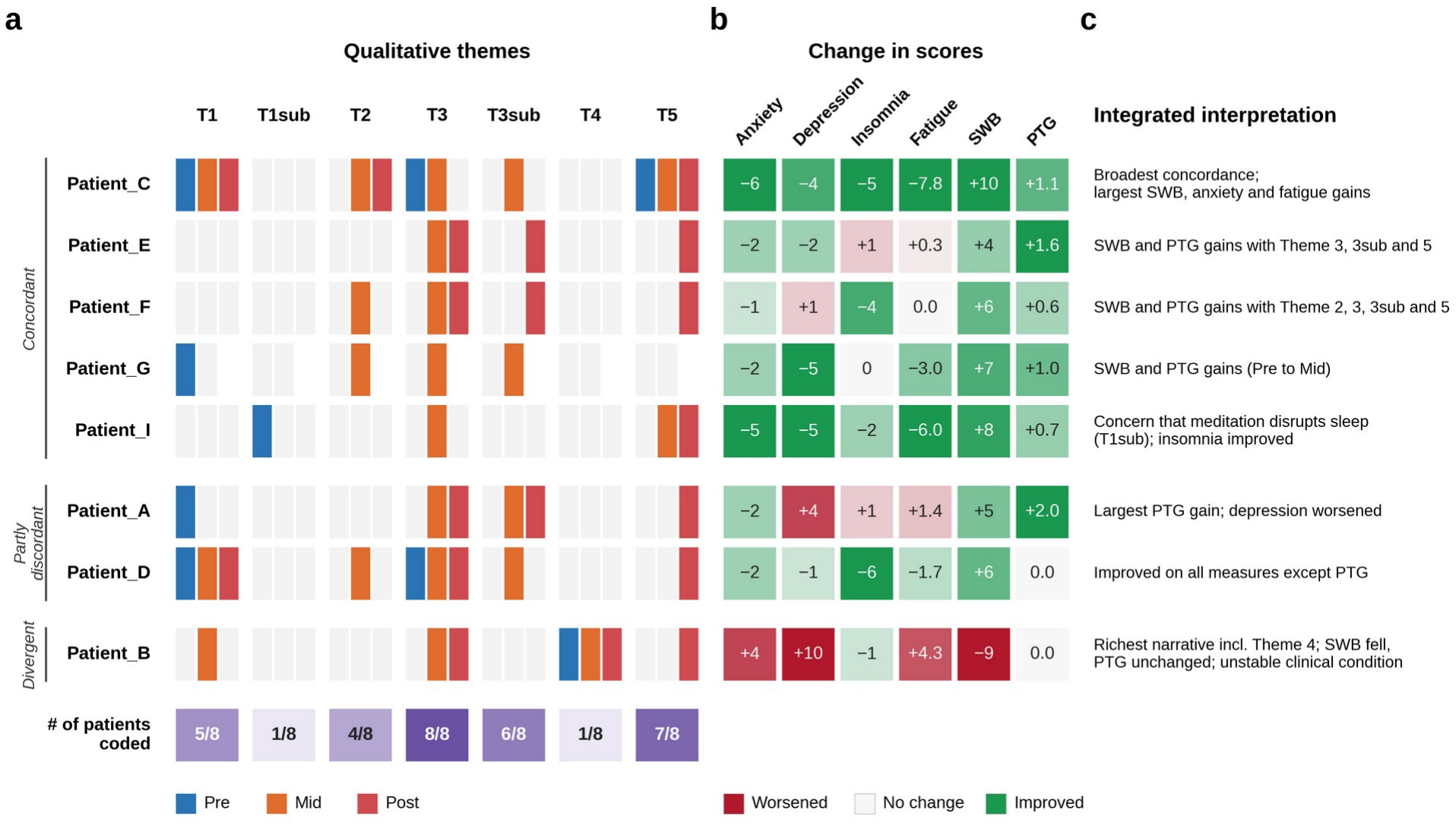
Joint display of qualitative themes and questionnaire score changes by participant. (a) Presence of qualitative themes (T1–T5) and subthemes (T1sub and T3sub); each cell is divided from left to right into Pre (blue), Mid (orange), and Post (red), and a colored segment indicates at least one coded passage for that theme and time point (light gray, none). The bottom row shows the number of participants coded to each theme at any time point, out of 8; darker purple indicates more participants. (b) Change in questionnaire scores from Pre to Post (Pre to Mid for Patient_G, who discontinued after the Mid assessment). Cell color indicates the direction and magnitude of change, with green indicating a favorable change (a decrease in anxiety, depression, insomnia, or fatigue scores, or an increase in SWB or PTG scores) and red indicating an unfavorable change. (c) Summary of the integrated interpretation for each participant. Rows represent the 8 participants who provided both questionnaire and free-text data, grouped by the pattern of quantitative-qualitative correspondence. Groupings reflect exploratory, case-level interpretations of correspondence between participants’ accounts and questionnaire changes. T1: uncontrollable anxiety and rumination; T1sub: concern about the practice of meditation itself; T2: relaxation through attention to breath and body; T3: disengagement from rumination and the attainment of calmness; T3sub: the observing self and acceptance; T4: existential and transcendent awareness beyond life and death; T5: internalization as a source of psychological support, extending into daily life.

#### Theme 1: Uncontrollable Anxiety and Rumination

At Pre, comments related to anxiety, fear, and rumination were observed in 4 of the 8 participants. Patient_G stated, “I feel anxious about the worsening side effects of chemotherapy” (Pre, Q1). Patient_A described being “tired of confronting the sudden fear and memories of unpleasant past events that come to mind” (Pre, Q1), and, regarding the hospital’s handling of care, remarked, “I sometimes feel as though I am being carried on a conveyor belt toward the end” (Pre, Q1), adding self-critically, “I feel ashamed that I imagine such terrible things” (Pre, Q1). Patient_D noted that, compared with the time of diagnosis and before surgery for colorectal cancer, “I now think about my illness less, and my mind has become calmer” (Pre, Q1), while also stating, “I sometimes feel a sudden anxiety about the future” (Pre, Q1).

In addition, Patient_C stated at Pre that “anxiety about side effects was substantial at the start of treatment” (Pre, Q1), indicating the presence of anxiety prior to program participation. In a retrospective account at Post, the same participant stated, “After being diagnosed with cancer, for a while I felt anxiety, fear, and a sense of loss, wondering whether my life was coming to an end” (Post, Q1), indicating that this participant had experienced profound existential fear in the early period of diagnosis and treatment.

A related but more specific concern, identified in 1 of the 8 participants and labeled Theme 1sub (concern about the practice of meditation itself), was raised by Patient_I at Pre: “Since I sometimes wake up in the middle of the night when I meditate, I feel anxious about whether I can continue” (Pre, Q1). Unlike the broader anxiety and rumination captured in Theme 1, this narrative reflected concern about the practice of meditation itself rather than about the illness or its treatment. The background to this concern and this participant’s subsequent trajectory are discussed under Integrating Quantitative and Qualitative Findings at the Individual Level.

#### Theme 2: Relaxation Through Attention to Breath and Body

Narratives related to breath and physical relaxation were observed in 4 of the 8 participants. At Mid, Patient_C described having learned the importance of focusing attention on the inflow and outflow of breath (Mid, Q2), and at Post reported specific physical changes: “After meditating, my mind and body feel more relaxed, and I feel I fall asleep more easily,” and “I have become able to sleep 7–8 hours” (Post, Q2). Patient_G stated, “Meditating seems to ease my tension” (Mid, Q1). Patient_F similarly stated, “I felt that I was getting a kind of rest different from sleep” (Mid, Q2). Patient_D reported that “bringing attention to my breath and taking deep breaths has become more common in daily life” (Mid, Q2), indicating that awareness of breathing had become embedded in daily life.

#### Theme 3: Disengagement From Rumination and the Attainment of Calmness

This was the only theme identified in the narratives of all 8 participants, representing the most widely shared change observed in this study. At Mid, Patient_D stated, “The time I spend thinking about my illness has decreased further” (Mid, Q1), and at Post reported, “I no longer think much about my illness,” except when physical symptoms were present (Post, Q1). At Mid, Patient_A stated, “There are still times when my mental state is poor, but I no longer dwell on it as much” (Mid, Q1), and at Post reported having “become able to regain a sense of calm” (Post, Q2). Patient_G stated, “There is no point in feeling impatient or worrying about things that cannot be helped” (Mid, Q2), and Patient_B remarked, “I have come to hear an inner voice telling me there is no need to be troubled” (Mid, Q2). Patient_E reported, “I have become able to spend my days more calmly than before” (Mid, Q1), and Patient_F reported, “The unexplained sense of anxiety has felt somewhat eased” (Mid, Q1).

Within this theme, a more specific subtheme (Theme 3sub: the observing self and acceptance) was identified in 6 of the 8 participants (Patient_A, Patient_C, Patient_D, Patient_E, Patient_F, and Patient_G). It comprised narratives in which participants described learning to observe their own minds nonjudgmentally and to accept them as they are. These accounts overlap with aspects of metacognitive awareness and decentering [36,37], as well as acceptance and attentional reorientation, although these processes were not directly measured. For example, Patient_D stated, “I feel I have become able to pause my thinking and bring my thoughts back to the present moment” (Mid, Q2); Patient_A described having come to know “another version of myself within, one calm enough to accept and embrace everything” (Post, Q2); and Patient_C stated, “I have become able to accept myself as I am” (Mid, Q1). These accounts suggest changes in participants’ relationship with their own thoughts and emotions, without establishing a specific underlying process.

#### Theme 4: Existential and Transcendent Awareness Beyond Life and Death

In contrast, this theme appeared in the narratives of only 1 of the 8 participants (Patient_B). At Mid, Patient_B described having come to hear an inner voice conveying that “I am being sustained by a great life force that runs from the center of the earth to the edge of the universe” (Mid, Q2), and stated, “I feel I have arrived at my own answer to why people are born and die” (Mid, Q1). At Post, the participant stated, “I sometimes feel a sense of reassurance, as though I have become one with the mysterious power of the great universe” (Post, Q2). Because this theme was identified in only a single participant, it should not be interpreted as a generalizable change; nevertheless, it is valuable as an individual case illustrating an existential experience reported during participation in Clinical Meditation.

#### Theme 5: Internalization as a Source of Psychological Support, Extending Into Daily Life

Descriptions in which meditation had become embedded in daily life and served as a source of psychological support, with effects extending to other areas of life such as concentration, relationships, and gratitude, were observed in all 7 participants who provided Post responses (Patient_G had discontinued after the Mid assessment). Patient_D described meditation as “something like a protective charm for the mind” (Post, Q2), Patient_C stated, “Meditation seems to have become my tranquilizer” (Post, Q2), and Patient_F remarked, “Knowing that I can always find calm through meditation has become a source of psychological support for me” (Post, Q2).

These accounts suggest that meditation had come to function as an inner resource rather than merely a learned technique. Others described effects extending beyond the practice itself: Patient_E reported, “I have become able to keep my mind calm, and my concentration has increased” (Post, Q2), and Patient_I stated, “I am truly grateful to the research team for creating this opportunity; I will do my best to keep it up on my own” (Post, Q2), expressing an intention to continue practicing independently. In addition, Patient_A stated, “I have become better than before at offering my own solutions to conflicts involving people close to me, in ways that do not burden them” (Post, Q1), suggesting a perceived change in interpersonal coping during the intervention period. Even while hospitalized because of worsening illness, Patient_B stated, “I was grateful that I could meditate even while in bed” (Post, Q1), illustrating that meditation could serve as a psychological resource beyond the context of daily routine.

Taken together, the 5 themes summarize participants’ reported experiences during the Clinical Meditation intervention. Comments related to uncontrollable anxiety and rumination (Theme 1) were most common at Pre, and relaxation via attention to breath and body (Theme 2) was described by 4 participants, while disengagement from rumination and the attainment of calmness (Theme 3) were described by all 8 participants over the course of the intervention. Existential and transcendent awareness beyond life and death (Theme 4), by contrast, was identified in only a single participant and should be regarded as an individual, deviant-case observation rather than a shared feature of the sample. At Post, 7 participants described meditation as a source of psychological support in daily life (Theme 5).

### Integrating Quantitative and Qualitative Findings at the Individual Level

Among the 8 participants who provided both quantitative and qualitative data, most showed engagement with a broad range of qualitative themes—particularly Theme 3 (disengagement and calmness) and Theme 5 (internalization as psychological support)—alongside increases in SWB and/or PTG scores (Figure 4). The following cases illustrate this correspondence and its limits.

Patient_C showed the broadest concordance between the two data sources: the largest increase in SWB (28 to 38) and the largest decreases in anxiety (−6) and fatigue (−7.8) in the sample were accompanied by narratives spanning nearly every qualitative theme, including an explicit account of improved sleep (Theme 2; “I have become able to sleep 7–8 hours”) that corresponded to this participant’s improvement in insomnia (−5). Patient_E, Patient_F, and Patient_G showed a similar, if more modest, concordant pattern, with increases in both SWB and PTG accompanying narratives centered on Theme 3 and Theme 3sub (for Patient_G, who discontinued after the Mid assessment, Pre-to-Mid changes).

Another example was Patient_I, whose concern at Pre that meditation itself might disrupt sleep (Theme 1sub) was, according to his remarks during the face-to-face group session, rooted in a previous unsuccessful experience with a different meditation program. Contrary to his initial concern, this participant showed a Pre-to-Post decrease in insomnia (−2), together with a marked decrease in anxiety (−5, from the highest Pre score in the sample) and narratives of disengagement from rumination (Theme 3) and internalization of meditation as an ongoing resource (Theme 5). He also remarked in an online group session that he had been breathing incorrectly in that earlier practice and had learned the correct way to breathe through the present program. Together, these observations suggest that the guidance provided in this program may have helped him acquire an appropriate meditation technique, including breathing, although this was not directly assessed.

Correspondence was not, however, uniform across measures within individual participants. Patient_A showed the largest increase in PTG in the sample (0.7 to 2.7) together with an increase in SWB (9 to 14), both from the lowest Pre scores in the sample, alongside narratives moving from uncontrollable anxiety and rumination at Pre (Theme 1) to self-acceptance by Post (Theme 3 and Theme 3sub; “another version of myself within, one calm enough to accept and embrace everything”); yet depression worsened (+4), and insomnia and fatigue worsened slightly (+1 and +1.4, respectively). Conversely, Patient_D showed favorable changes on every measure except PTG, which was unchanged (3.6 to 3.6), including the largest decrease in insomnia in the sample (−6), alongside narratives of disengagement from illness-related rumination (Theme 3; “I no longer think much about my illness”).

Patient_B represented a more pronounced divergence: despite the richest qualitative narrative in the study, including the only substantial existential and transcendent content (Theme 4), PTG did not increase from Pre to Post (2.8 to 2.8, after peaking at 4.4 at Mid) and SWB declined markedly (37 to 28), coinciding with an unstable clinical status, including hospitalization for multiple complications, around the Post assessment (see Theme 5).

In aggregate, these participant-level comparisons suggest that quantitative and qualitative changes generally moved in the same direction for most participants, particularly with respect to Theme 3, Theme 5, and increases in PTG or SWB, but that the correspondence was neither uniform across outcome measures within a given participant (Patient_A, Patient_D) nor guaranteed even when qualitative content was rich (Patient_B).

## Discussion

### Principal Findings

In this mixed-methods pilot study, a Clinical Meditation guidance intervention was delivered to 11 patients with cancer who were receiving or scheduled to receive pharmacotherapy at recruitment. Of the 11 enrolled participants, 9 (81.8%) completed the Mid assessment and 8 (72.7%) completed the Post assessment, and attendance at the online group sessions ranged from 45% to 73%. Self-reported meditation practice frequency, including guided sessions and home practice, decreased from a median of 5–6 days per week in the structured first month to 3–4 days per week in the self-directed second month.

SWB and PTG were the only outcomes showing significant overall change: SWB increased by 4 weeks but declined by 8 weeks, whereas PTG increased and remained above baseline at 8 weeks. Anxiety, depression, insomnia, and fatigue showed no significant change. In the qualitative analysis, comments related to uncontrollable anxiety and rumination were most common before the intervention, whereas 4 participants later described relaxation through attention to breath and body and all 8 described disengagement from rumination. At Post, 7 participants described meditation as an ongoing source of psychological support. At the individual level, integration of questionnaire findings and participants’ accounts revealed both correspondence and divergence.

### Comparison With Prior Work

With respect to feasibility, the 8-week attrition rate in our study (27.3%) was numerically similar to the average of 25.3% reported in a systematic review of digitally delivered mindfulness-based programs for patients with cancer, although differences in populations and attrition definitions limit direct comparison [38]. All 11 participants attended the initial face-to-face session, and 8 of the 9 participants (88.9%) who continued beyond it completed the 8-week assessment. In a qualitative study of a mindfulness-based group program in primary care, participants reported that having first met in person helped them engage in subsequent videoconference sessions [39].

Initial in-person contact followed by ongoing support may therefore have helped sustain participation in our study. However, 2 participants discontinued after the initial session, and the transition from face-to-face to online delivery may have been a barrier to participation. One of these participants had an unstable clinical condition, and the reasons for discontinuation were not confirmed for either. Future studies should identify barriers during this transition and compare hybrid and fully online formats to clarify the advantages and challenges of hybrid delivery.

Among the 9 participants who continued beyond the initial session, mean attendance at the 3 online group sessions was 74.1% (20/27 session attendances; 55.6%, 77.8%, and 88.9% for each session), lower than the mean attendance of 90% reported in a 4-week online Mindfulness-Based Cancer Recovery (MBCR) trial [40]. In addition to live group instruction, that trial provided video recordings of sessions to absent participants, an online messaging group for sharing materials and communication, daily practice logging via a smartphone app, and training in the use of the videoconferencing system. Although the populations, programs, and calculation methods differed, and the effect of each strategy was not examined individually, these strategies may be useful for supporting participation. Why participants missed sessions, and whether such support improves session attendance and continued practice, remain to be examined.

In the self-directed second month, median practice frequency declined from 5–6 days per week to 3–4 days per week, even though 1 individual online session was provided. Because practice frequency included session attendance and home practice changed from prescribed to voluntary, this decline partly reflects changes in the program structure. The role of individual support in sustaining practice also remains unclear. In a retrospective study of online MBSR, adding brief individual interviews to weekly group classes was associated with greater improvements in psychological well-being, but home practice time was shorter than without interviews [41].

Individual sessions may therefore contribute to outcomes through mechanisms other than increasing practice. Masheder et al identified factors relevant to sustaining practice, including self-efficacy, experiencing the rewards of practice, social support, and the relationship with the teacher [42]. On this basis, future studies should evaluate several strategies for sustaining practice, along with the time and staff they require, while considering the burden on both participants and instructors. These include adding brief online group sessions in the second month, adjusting the timing and frequency of individual support, providing regular feedback, and developing practice plans tailored to participants’ daily lives.

Regarding psychological outcomes, the transient increase in SWB in our study contrasts with the sustained increase in PTG. Previous trials suggest that SWB benefits may vary over time. Park et al reported improvements that were maintained 4 weeks after completion of MBCT [11], whereas the improvement in meaning and peace reported by Bower et al was no longer significantly greater than that in controls at 3-month follow-up [43]. Similarly, Würtzen et al found a between-group difference in SWB at 6 months but not at 12 months, with no significant effect in the longitudinal mixed-effects analysis [44]. In our study, the decline in SWB coincided with reduced practice during the less structured second month. Nevertheless, 7 of the 8 participants remained above baseline at 8 weeks; the only participant below baseline (Patient_B) was hospitalized for complications around the Post assessment, which likely contributed to the nonsignificant Pre-to-Post difference in this small sample. Whether continued guidance could help sustain practice and SWB warrants further study.

The PTG findings can also be considered alongside trials of mindfulness-based interventions. Carlson et al found greater increases in FACIT-Sp and Posttraumatic Growth Inventory (PTGI) total scores with MBCR than with supportive expressive group therapy, with PTGI total scores continuing to increase relative to the comparison group during follow-up [45]. Zhu et al also reported higher postintervention PTGI scores following an 8-week MBSR program among women receiving chemotherapy for breast cancer [46]. These findings provide context for the PTG changes observed in our study, although differences in intervention content, populations, and study design limit direct comparison. Importantly, PTG can also develop without any psychological intervention: in observational studies of patients with breast cancer, PTG scores increased over the months following diagnosis [47,48], with most of the increase occurring within the first few months [47]. Unlike the trials above, our pilot study had no control group, so part of the observed increase in PTG may reflect this natural course, and we cannot determine whether Clinical Meditation promoted PTG beyond it.

The qualitative findings offer a complementary perspective on these outcomes. Against the background of anxiety and rumination described in Theme 1, attention to breathing and bodily relaxation in Theme 2 may have provided an initial respite, allowing participants to gain distance from rumination, as described in Theme 3, and, as captured in Theme 3sub, to observe and accept their thoughts and feelings as they are. This shift may have opened opportunities for existential reflection, reported by one participant in Theme 4, and for meditation to become an ongoing source of support, as reflected in Theme 5. This pattern of themes is consistent with PTG theory and mindfulness-to-meaning theory. PTG theory proposes that growth may emerge as individuals reconsider beliefs disrupted by a major life crisis and reconstruct meaning through deliberate reflection [8]. Similarly, mindfulness-to-meaning theory describes how decentering—viewing thoughts and feelings as passing experiences—may broaden awareness and support more flexible interpretations of adversity [49]. These themes suggest a possible pathway through which relief from distress and greater distance from rumination could support meaning-making and changes in SWB and PTG [50].

Related experiences have also been reported in qualitative studies of mindfulness in cancer care. Ando et al [51] identified adaptive coping, positive meaning, personal growth, and spirituality in Japanese patients with cancer following mindfulness-based meditation. Similarly, Mackenzie et al [52] identified personal growth and spirituality among the major themes in interviews and a focus group with cancer patients who had completed MBSR and continued attending meditation sessions. These findings provide context for interpreting our participants’ accounts of existential reflection and meditation as a source of psychological support. In a recent mixed-methods study of online MBCR, Gaignard et al [19] assessed SWB and PTG alongside participants’ experiences. Qualitative interviews revealed enhanced well-being, skills acquisition, and engagement with cancer-related memories, which some participants found challenging. Such accounts help illuminate how participants understand changes in well-being, coping, and their relationship with illness.

In our study, integrating participants’ own accounts with their questionnaire scores illustrated both correspondence and divergence between the two data sources. For Patient_C, accounts of relaxation and improved sleep accompanied an increase in SWB and decreases in several symptom scores. Patient_A described greater self-acceptance alongside increased PTG and SWB, although depression, insomnia, and fatigue scores increased. Conversely, Patient_B’s existential reflections accompanied unchanged PTG and declining SWB during a period of clinical instability. These patterns show that perceived benefits may coexist with difficulties in other domains and that existential reflection does not necessarily coincide with measured growth or improved well-being. Qualitative accounts therefore added context to the questionnaire findings by describing what participants experienced and how meditation featured in their daily lives.

### Limitations

This study has several limitations. Regarding feasibility, no criteria for recruitment, retention, or adherence were prespecified, so the observed engagement provides descriptive information rather than a definitive assessment of feasibility. Participants who consented may have had a preexisting interest in meditation. Because the reasons for the 2 discontinuations after the initial session were not confirmed, the remaining sample may represent a subgroup more willing or able to continue. Future studies should define feasibility criteria in advance, drawing on established frameworks [53].

For the questionnaire outcomes, the quantitative sample was small (n=9), and the single-arm design precludes attributing the observed changes to the intervention. Natural course, regression to the mean, and repeated-testing effects cannot be ruled out. Six outcome measures were assessed at 3 time points without a prespecified primary outcome; despite adjustment for multiplicity, the significant findings require confirmation. Differences in cancer type, stage, and treatment could not be accounted for, and the 8-week follow-up was too short to assess longer-term trajectories, particularly the persistence of changes in SWB and PTG.

Regarding the qualitative findings, the data were limited to free-text responses rather than semi-structured interviews, and some responses were brief, so experiences not spontaneously described may have been missed. With 8 participants, theme saturation was not confirmed, and Theme 4 was based on a single participant. Social desirability bias may have influenced positive narratives such as those in Theme 5, while AN’s involvement in recruitment and group sessions may also have influenced participants’ responses and the interpretation of the qualitative data. The integration of questionnaire scores and participants’ accounts was exploratory and intended to aid interpretation of individual experiences. Although the themes offer possible interpretations of participants’ experiences in relation to PTG theory and mindfulness-to-meaning theory, the present data do not establish a temporal sequence among the themes or demonstrate that these experiences accounted for changes in SWB or PTG. Accordingly, the potential mechanism suggested by the qualitative findings warrants further investigation using measures of rumination, such as the Event-Related Rumination Inventory [54], and decentering, such as the Experiences Questionnaire [37].

### Conclusions

This mixed-methods pilot study suggests that Clinical Meditation may be feasible and acceptable for patients with cancer. Combining an initial face-to-face session with brief online group and individual sessions may offer a practical approach to providing support during cancer treatment, although the decline in meditation practice frequency highlights challenges in sustaining engagement. Exploratory increases in SWB and PTG, together with participants’ accounts of gaining distance from ruminative thoughts and finding support through meditation, warrant further investigation. Larger controlled studies with longer follow-up are needed to further assess feasibility and evaluate psychological outcomes. Comparisons with established MBIs may help clarify the distinctive features of Clinical Meditation, while more in-depth qualitative research could clarify how patients experience and respond to the intervention.

## Supporting information

Multimedia Appendix 1: Clinical Meditation Protocol

Multimedia Appendix 2: Supplementary Figures and Tables

Multimedia Appendix 3: SRQR Checklist

Multimedia Appendix 4: Qualitative Codebook

## Acknowledgments

We would like to express our sincere gratitude to all the patients who participated in this study. We also thank Keiko Kusaka for reviewing the manuscript and providing helpful comments. LLMs were used as primary tools for statistical analysis, figure generation, qualitative analysis, literature checking, and language editing of the manuscript. AN reviewed and verified all LLM-generated outputs and made the final analytical and interpretive decisions. AN takes responsibility for the accuracy and integrity of the reported work.

## Data Availability

The quantitative data generated and analyzed during this study are available from the corresponding author on reasonable request. Full individual free-text responses are not publicly available to protect participant privacy. The qualitative codebook and theme structure, including deidentified English translations of all coded passages, are provided in Multimedia Appendix 4.

## Authors’ Contributions

AN contributed to study conceptualization and design, participant recruitment, overall data analysis, and drafting of the manuscript. TK contributed to the design of the Clinical Meditation guidance program and delivery of the meditation intervention. KB and GF contributed to participant recruitment. NI contributed to qualitative analysis. DO contributed to the design of the Clinical Meditation guidance program and supervision of intervention delivery. DS contributed to study conceptualization and design. NB contributed to study conceptualization and design and participant recruitment. All authors reviewed and approved the final manuscript.

## Abbreviations

BFI: Brief Fatigue Inventory
FACIT-Sp: Functional Assessment of Chronic Illness Therapy-Spiritual
HADS: Hospital Anxiety and Depression Scale
ISI: Insomnia Severity Index
LLM: large language model
MBCR: Mindfulness-Based Cancer Recovery
MBCT: mindfulness-based cognitive therapy
MBI: mindfulness-based intervention
MBSR: mindfulness-based stress reduction
PTG: posttraumatic growth
PTGI: Posttraumatic Growth Inventory
PTGI-X-SF-J: Japanese version of the Short Form of the Expanded Version of the Posttraumatic Growth Inventory
QOL: quality of life
SRQR: Standards for Reporting Qualitative Research
SWB: spiritual well-being

**Multimedia Appendix 1**

Clinical Meditation background and intervention protocol.

**Multimedia Appendix 2**

Supplementary figures (Figures S1–S6) and tables (Tables S1 and S2).

**Multimedia Appendix 3**

Standards for Reporting Qualitative Research (SRQR) checklist.

**Multimedia Appendix 4**

Qualitative codebook, including the thematic structure and English translations of all coded passages.

