## Supplementary material for "Feasibility and Psychological Outcomes of a Clinical Meditation Intervention for Patients with Cancer: A Mixed-Methods Pilot Study": Multimedia Appendix 1: Clinical Meditation Protocol

### Multimedia Appendix 1. Clinical Meditation Intervention Protocol

**Clinical Meditation: Background and Intervention Protocol**

**Overview of Clinical Meditation**

Clinical Meditation (Rinsho Meiso-ho) was developed by Daien Ohshita, Elder of Senkoji Temple and director of the Clinical Meditation Education and Research Institute. After training in meditation at Koyasan in the Shingon esoteric Buddhist tradition and in Sri Lanka in the Theravada Buddhist tradition, Ohshita investigated the clinical applications of meditation at Kyoto University's Kokoro Research Center. Drawing on a review of meditation practices from around the world, he cross-referenced their functions, purposes, and applications and integrated them with contemporary medical and psychological findings and the distinctive features of Buddhist meditation. This work culminated in the systematization of Clinical Meditation.

In approximately 10 years since its development, Clinical Meditation has been used in a wide range of clinical and community settings, including end-of-life care, psychological support in disaster-affected areas, and stress-management programs for corporate employees and healthcare and long-term care professionals.

Within the Clinical Meditation framework, meditation techniques and their associated states of consciousness are classified into four groups:

①Loosen Meditation (Yurumeru Meiso): A form of meditation intended to relax and focus the mind and body by deliberately harmonizing the rhythms of breathing and bodily movement.

②Observation Meditation (Mitsumeru Meiso): A form of meditation in which practitioners build on the stable state of consciousness attained through Loosen Meditation and objectively observe themselves and others, thereby deepening awareness and insight.

③Energizing Meditation (Takameru Meiso): A form of meditation intended to enhance the capacities and potential of the mind and body through the deliberate use of the five senses and bodily sensations.

④Unifying Meditation (Yudaneru Meiso): A state of consciousness that develops from Energizing Meditation, in which practitioners relinquish attachment to the ego and experience a sense of unity or oneness with a transcendent presence, variously described as God, Buddha, or "something great."

Within this classification, mindfulness meditation corresponds primarily to Loosen Meditation and Observation Meditation. By being systematized without emphasizing its religious origins, mindfulness meditation has achieved widespread adoption in fields such as healthcare and education. However, some have argued that its emphasis on practical benefits, including stress reduction and enhanced performance, has come at the expense of adequate attention to the religious and existential dimensions inherent in Buddhist meditation.

By contrast, although Clinical Meditation can likewise be practiced by anyone, irrespective of faith or religious affiliation, it explicitly incorporates elements of Buddhist meditation intended to help practitioners find their own answers to the existential and spiritual questions that arise when confronting illness and death. Such questions include "Why do I live?" "What is the purpose of life?" and "What meaning do illness and death hold?" Clinical Meditation also encompasses a sense of unity with "something greater," a concept expressed not only in Buddhism but also in Christianity and other religious traditions.

Given these characteristics, Clinical Meditation may offer a stepwise approach to supporting patients experiencing spiritual pain associated with illness, including anxiety, inner conflict, and questions about the meaning of life. Through Loosen Meditation and Observation Meditation, practitioners observe their bodies, emotions, and thoughts as they are, thereby deepening self-understanding without turning away from their suffering. On this foundation, they progress to Energizing Meditation and Unifying Meditation, with the aim of developing a broader perspective that extends beyond the self and discovering renewed meaning and hope in life. This progressive approach may provide a practical framework for the spiritual care of patients experiencing spiritual pain.

**Meditation Program Used in This Study**

In this study, participants practiced meditation according to the following protocol. The program centered on Loosen Meditation, which is accessible even to people with no prior meditation experience, and incorporated elements of Observation Meditation and Energizing Meditation.

① Warm-up stretches (15 minutes in the face-to-face session; 5 min in the online sessions): Participants performed several stretches targeting different parts of the body, coordinating each movement with slow breathing.

② Posture adjustment (1 minute): Participants sat in a chair with their eyes gently closed, pelvis upright, and spine naturally extended. They rested both hands on their thighs or loosely clasped them in front of the abdomen and released tension from the shoulders and arms.

③ Deep breathing (2 minutes): Participants inhaled slowly through the nose, imagining the lungs and rib cage expanding fully, and exhaled through the mouth in a thin, prolonged stream. This cycle was repeated for approximately 10 breaths.

④ Body scan (7 minutes): Participants sequentially directed their attention to the following body regions, releasing tension with each exhalation: (1) the crown of the head; (2) the areas around the eyes and between the eyebrows; (3) the nose, mouth, and cheeks; (4) the neck and throat; (5) the left and right shoulders; (6) the left and right arms; (7) the chest, abdomen, and back; (8) the buttocks and groin; (9) the left and right legs; and (10) the body as a whole.

⑤ Breath-observation meditation (15 minutes): While breathing naturally, participants directed and sustained their attention on the breath. When thoughts or emotions arose and their attention wandered, they were instructed neither to reject nor suppress these experiences. Instead, after noticing the distraction, they gently redirected their attention to the breath.

⑥ Loving-kindness meditation (3 minutes): Participants directed feelings of loving-kindness and gratitude outward while slowly and silently reciting the following phrases:

*May I be well.*

*May I be happy.*

*May those dear to me be well.*

*May those dear to me be happy.*

*May all living beings be well.*

*May all living beings be happy.*

*May the weakened organs within me receive my gratitude and care, and may they gently move toward recovery.*

⑦ Closing (1 minute): Participants slowly opened their eyes, gently stretched their entire bodies, and concluded the meditation.

The in-person session and the first two online group sessions followed this protocol without step ⑥, whereas the third online group session and the individual online session included all steps. Each session ended with reflection on participants’ meditation experiences and current physical and psychological state, followed by questions and answers about the techniques. For home practice, participants used a shorter audio-guided sequence comprising only breathing exercises, a body scan, and breath-observation meditation and were asked to practice for 15 minutes on days without scheduled sessions during the first month. During the second month, participants who wished to continue were free to practice independently at their own discretion.
