## Supplementary material for "Feasibility and Psychological Outcomes of a Clinical Meditation Intervention for Patients with Cancer: A Mixed-Methods Pilot Study": Multimedia Appendix 2: Supplementary Figures and Tables

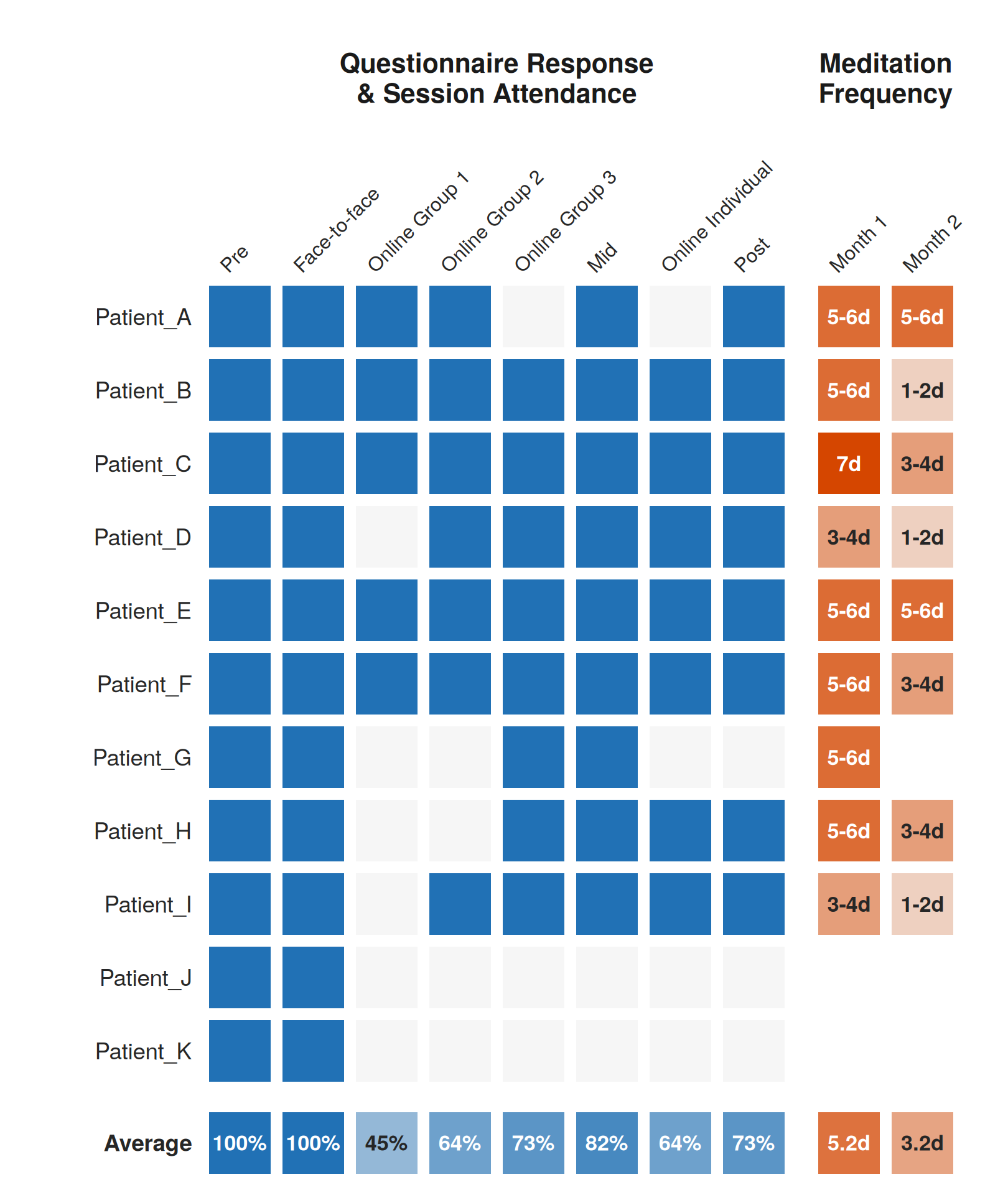

**Figure S1.** Individual session attendance and meditation practice frequency. Heat map of questionnaire completion and session attendance (blue; light gray indicates not completed or not attended) and home meditation practice frequency (orange scale, by days per week) for each of the 11 enrolled participants. The bottom row shows the group average.

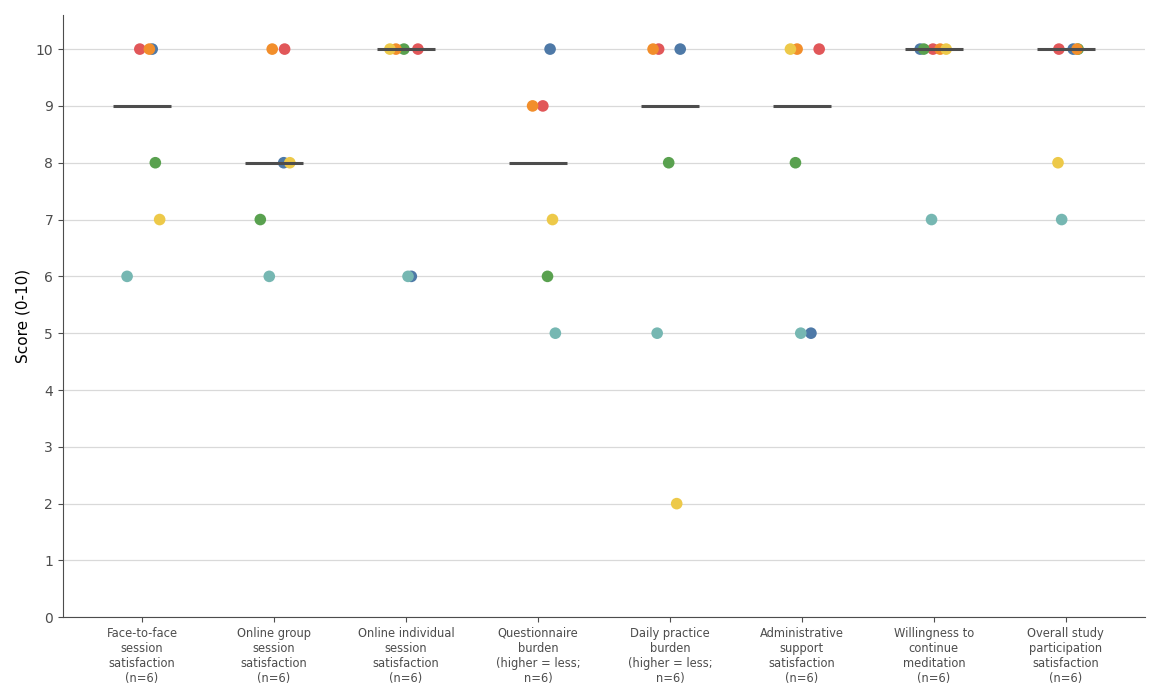

**Figure S2.** Dot plots show responses to an anonymous questionnaire assessing participants’ experiences of study participation, administered after completion of the Post assessment. Responses were obtained from six participants. Each colored dot represents an anonymous response and cannot be linked to an individual participant. Horizontal lines indicate the median.

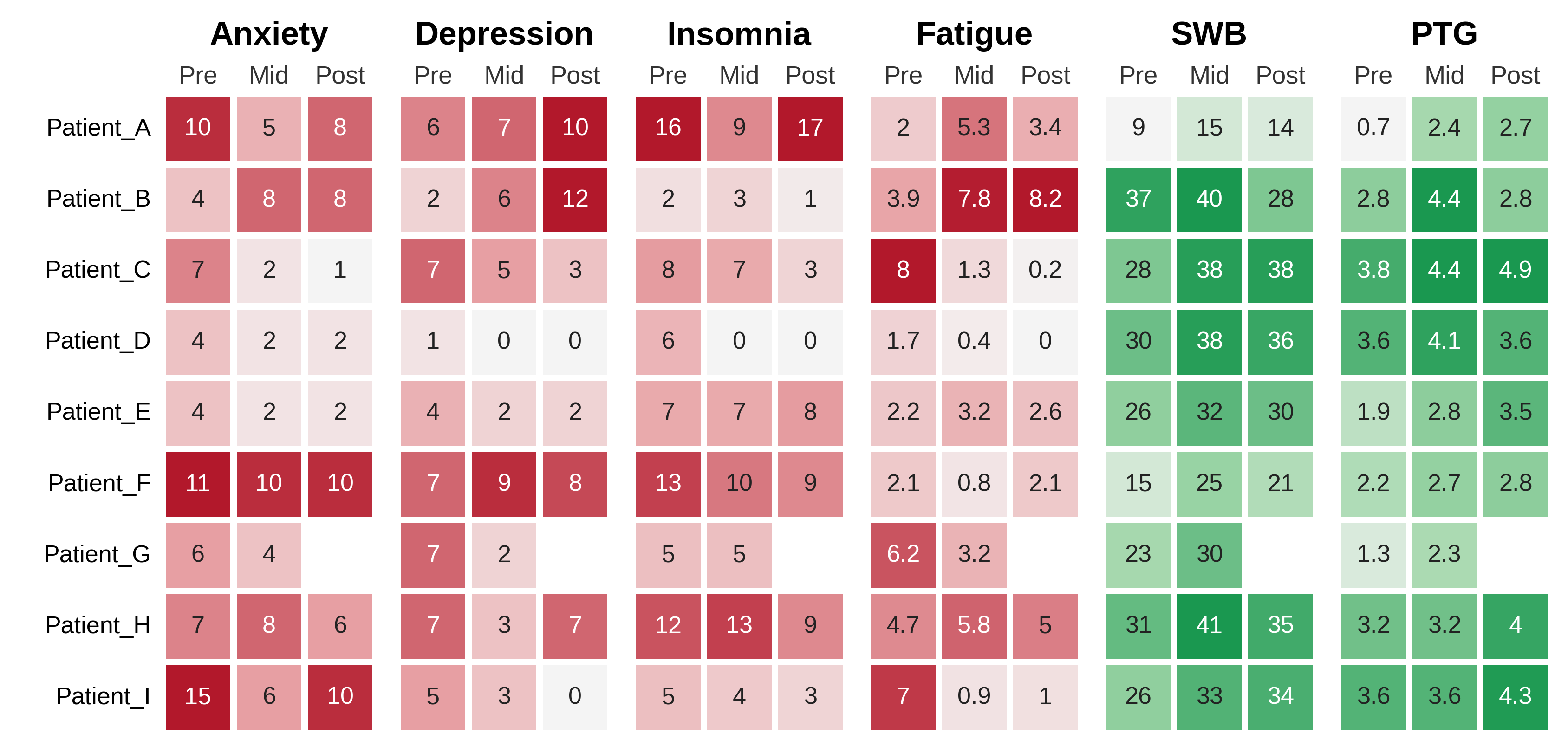

**Figure S3.** Individual participant questionnaire scores. Heat map of scores on the 6 outcome measures for each of 9 participants. Because higher scores indicate improvement for SWB and PTG but worsening for anxiety, depression, insomnia, and fatigue, the two groups are shown in different colors: green and red, respectively. Color intensity reflects the relative magnitude within each measure. Patient_G has no data at the Post time point.

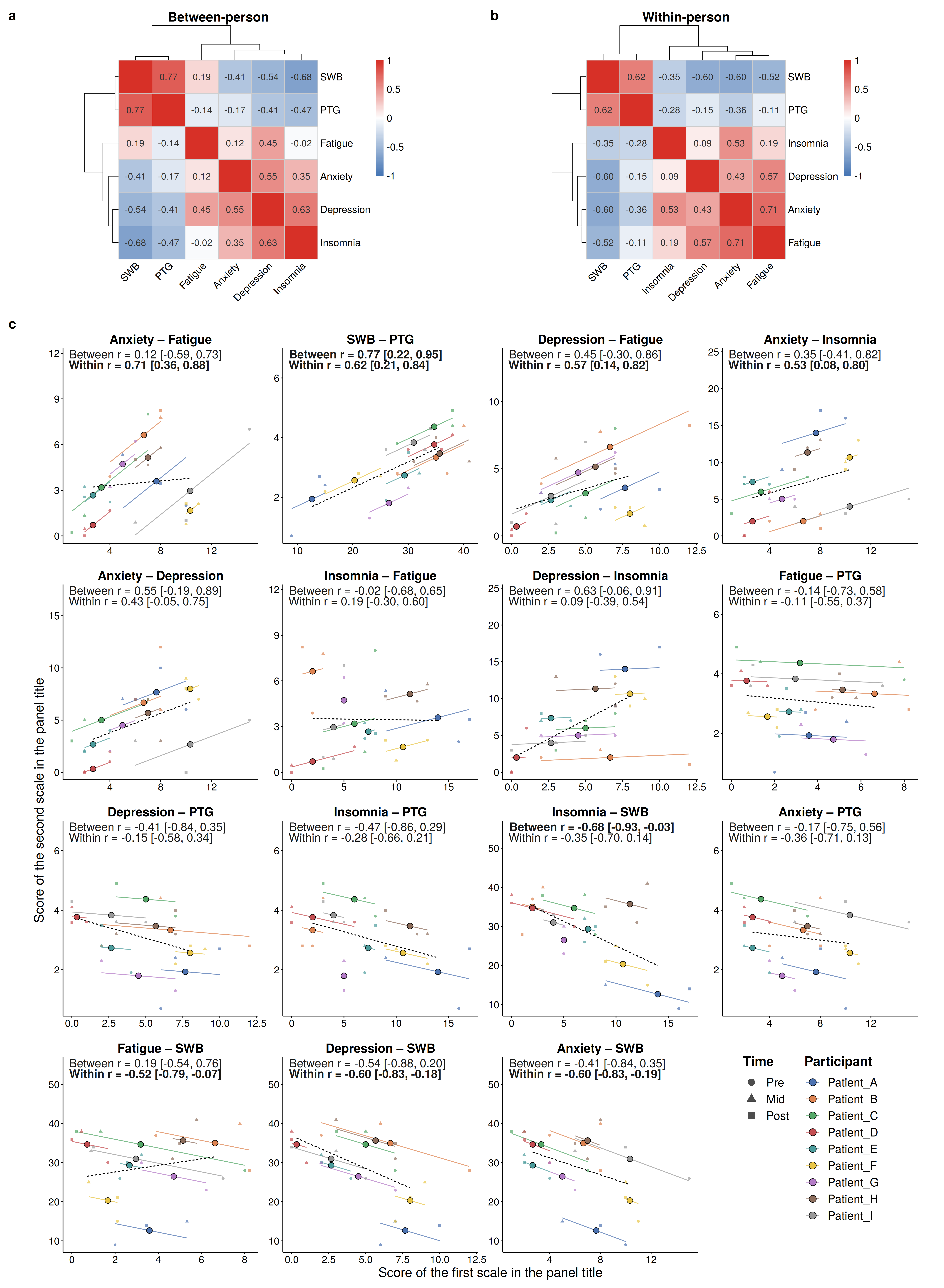

**Figure S4.** Associations among the 6 outcome measures. Heat maps show (a) between-person correlations (Pearson correlation of participant means across the 3 time points) and (b) within-person correlations (repeated-measures correlation; rmcorr) for all 15 pairwise combinations of the 6 outcome measures; rows and columns are ordered by Ward hierarchical clustering (distance=1 − r). (c) Scatter plots for each pairwise combination; small points show individual observations at each time point (circle=Pre, triangle=Mid, square=Post), colored by participant. Large circular points show each participant’s mean across the 3 time points. Colored lines show each participant’s within-person regression line; the dashed black line shows the between-person regression line. Each panel reports the between-person and within-person r with 95% CIs (Fisher z transformation); bold text indicates a 95% CI excluding zero.

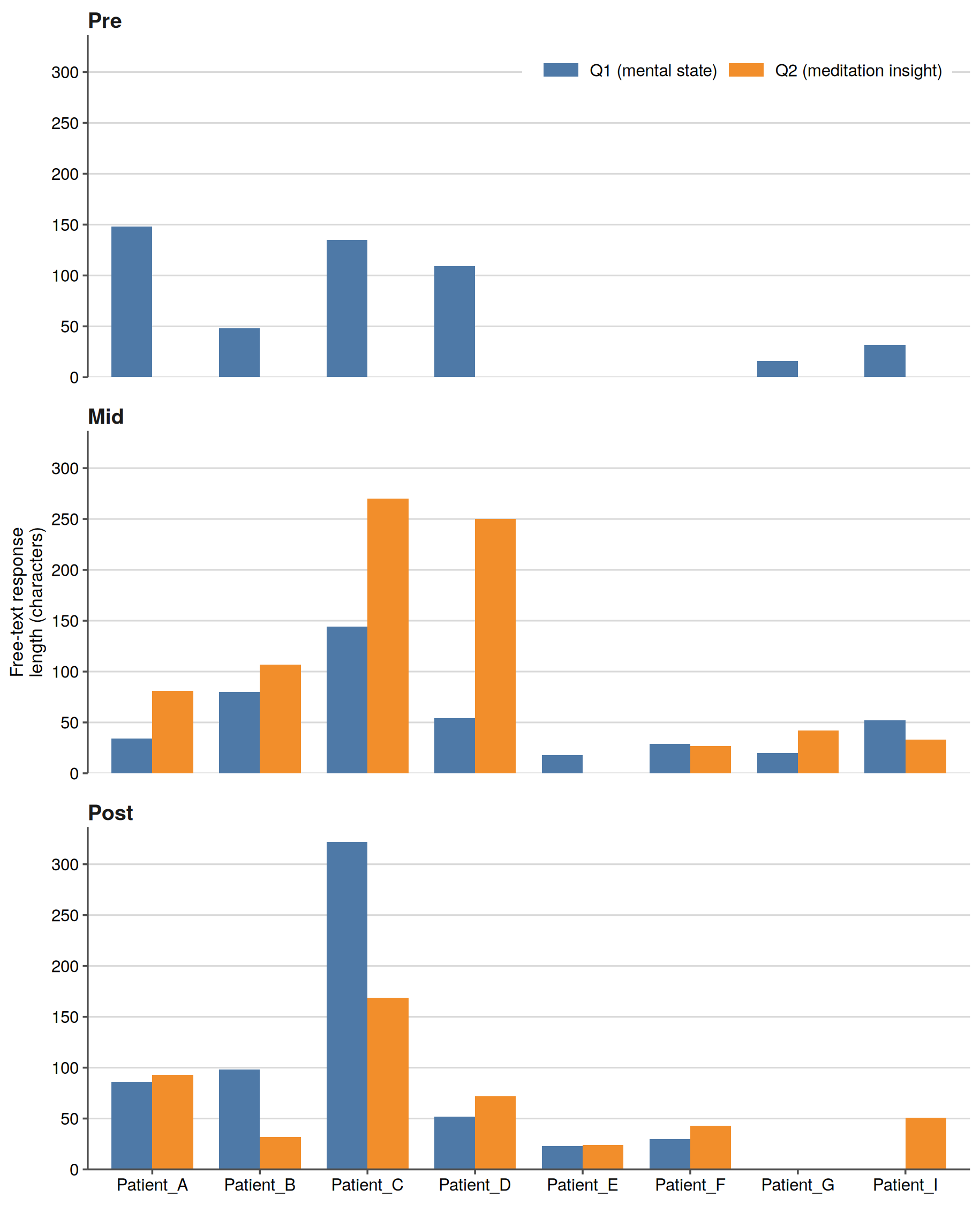

**Figure S5.** Length of free-text responses by participant, time point, and question. Bar chart showing the number of characters in each qualitative free-text response (Q1: current state of mind; Q2: insights or changes gained through meditation practice) for each of the 8 participants who provided free-text data, at each of the 3 time points (Pre, Mid, Post). An absent bar indicates that no response was collected for that participant, time point, and question (eg, Patient_G discontinued after the Mid assessment; Q2 was not administered at Pre).

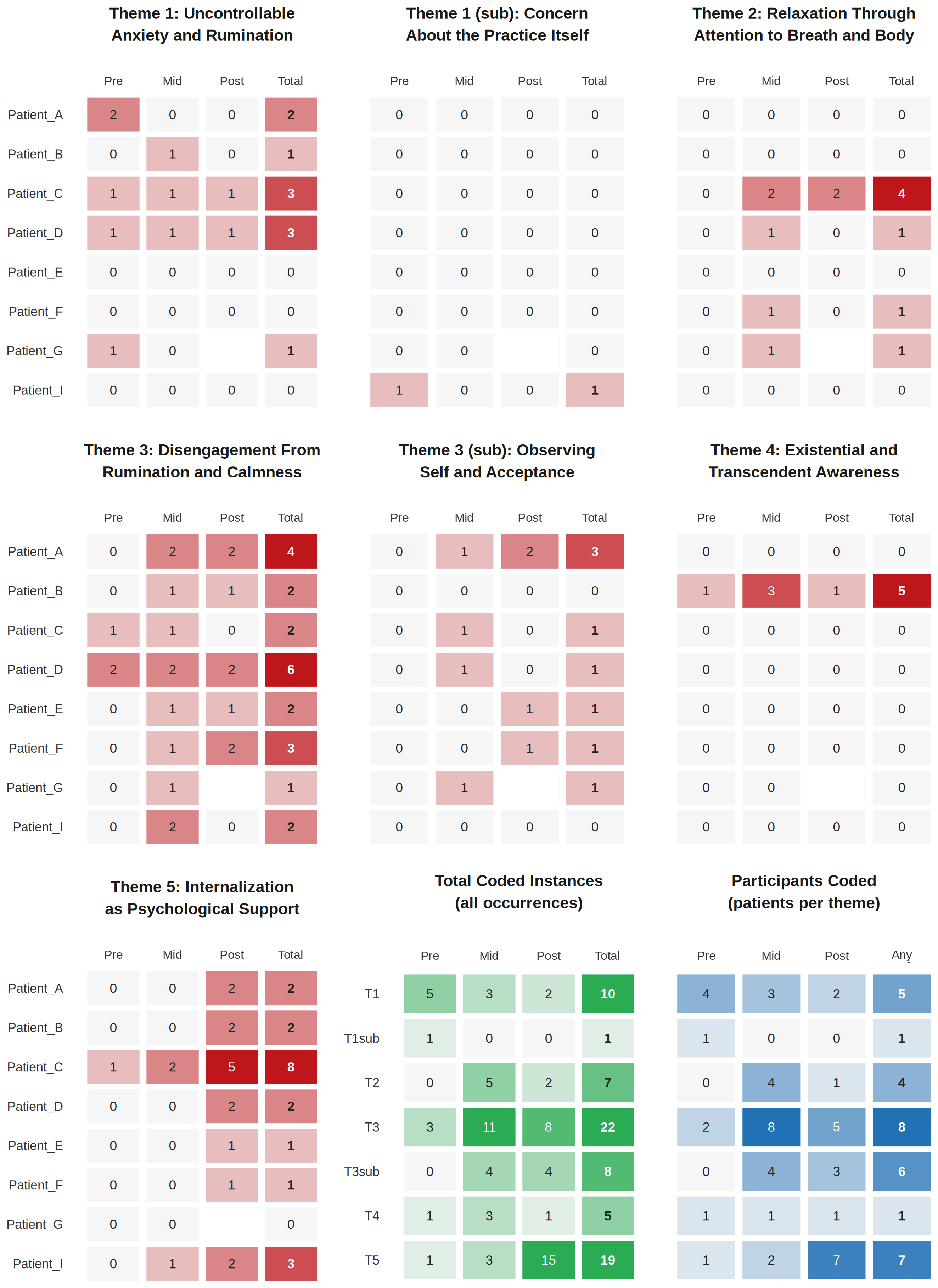

**Figure S6. Detailed coded-passage counts by qualitative theme, participant, and time point.** The 7 theme and subtheme panels show coded-passage counts for each of 8 participants at Pre, Mid, Post, and across all time points (Total). All 7 panels share a red scale, with counts of 4 or more shown in the darkest red. The green panel summarizes coded-passage counts across participants by theme and time point, with totals of 10 or more shown in the darkest green. The blue panel shows participant counts (0–8) for each theme at each time point and at any time point (Any). Patient_G discontinued after Mid. Figure 4 presents a summary of theme presence using the same coding data.

**Table S1. Friedman test results for questionnaire score changes.**

For each of the 6 outcome measures, overall change across the 3 time points (Pre, Mid, Post) was assessed using the Friedman test among the 8 participants with complete data at all 3 time points; P values were adjusted for the 6 outcome measures using the Benjamini-Hochberg procedure. SWB and PTG (bold) were the only 2 outcomes with a significant omnibus result.

| **Scale** | **Friedman χ²** | **P** | **Adjusted P** |
| --- | --- | --- | --- |
| Anxiety | 5.43 | 0.066 | 0.13 |
| Depression | 0.48 | 0.79 | 0.91 |
| Insomnia | 3.27 | 0.20 | 0.29 |
| Fatigue | 0.19 | 0.91 | 0.91 |
| **SWB** | **11.8** | **0.0027** | **0.016** |
| **PTG** | **8.86** | **0.012** | **0.036** |

**Table S2. Pairwise nonparametric effect sizes for questionnaire score changes.**

t1 and t2 denote the earlier and later of the two time points being compared in each row (e.g., for the Mid - Pre comparison, t1 = Pre and t2 = Mid). Medians and interquartile ranges (IQR) are shown at each time point. The Hodges-Lehmann estimate and the matched-pairs rank-biserial correlation (r) were both calculated as the change from t1 to t2 (t2 minus t1); the Hodges-Lehmann estimate is the median of all pairwise Walsh averages of the paired differences, with a distribution-free 95% CI derived from the same exact Wilcoxon signed-rank procedure used for the P value, and the matched-pairs rank-biserial correlation (r) is the nonparametric effect size consistent with the Wilcoxon signed-rank test. Given the small sample size, the exact (permutation-based) form of the Wilcoxon signed-rank test was used for the P value in every row, including the purely descriptive rows. For SWB and PTG, the 2 outcomes with a significant Friedman result in Table S1, the Adjusted P column reports the Shaffer-adjusted P value from post hoc testing within that outcome's 3 comparisons.

| **Scale** | **t1** | **t2** | **n** | **Median (IQR), t1** | **Median (IQR), t2** | **Hodges-Lehmann (95% CI)** | **Rank-biserial r** | **P** | **Adjusted P** |
| --- | --- | --- | --- | --- | --- | --- | --- | --- | --- |
| Anxiety | Pre | Mid | 9 | 7.0 (4.0-10) | 5.0 (2.0-8.0) | -2.0 (-5.5 to 1.0) | -0.67 | 0.098 | — |
| Anxiety | Mid | Post | 8 | 5.5 (2.0-8.0) | 7.0 (2.0-8.5) | 1.0 (-2.0 to 4.0) | 0.40 | 0.62 | — |
| Anxiety | Pre | Post | 8 | 7.0 (4.0-10) | 7.0 (2.0-8.5) | -2.0 (-4.0 to 1.0) | -0.67 | 0.11 | — |
| Depression | Pre | Mid | 9 | 6.0 (4.0-7.0) | 3.0 (2.0-6.0) | -1.5 (-3.5 to 1.5) | -0.40 | 0.36 | — |
| Depression | Mid | Post | 8 | 4.0 (2.8-6.2) | 5.0 (1.5-8.5) | 1.0 (-2.0 to 4.5) | 0.38 | 0.56 | — |
| Depression | Pre | Post | 8 | 5.5 (3.5-7.0) | 5.0 (1.5-8.5) | 0.0 (-4.5 to 7.0) | -0.071 | 0.94 | — |
| Insomnia | Pre | Mid | 9 | 7.0 (5.0-12) | 7.0 (4.0-9.0) | -2.3 (-5.0 to 0.0) | -0.64 | 0.16 | — |
| Insomnia | Mid | Post | 8 | 7.0 (3.8-9.2) | 5.5 (2.5-9.0) | -1.2 (-3.0 to 3.5) | -0.36 | 0.47 | — |
| Insomnia | Pre | Post | 8 | 7.5 (5.8-12) | 5.5 (2.5-9.0) | -2.5 (-5.0 to 0.0) | -0.78 | 0.055 | — |
| Fatigue | Pre | Mid | 9 | 3.9 (2.1-6.2) | 3.2 (0.90-5.3) | -1.1 (-4.0 to 2.2) | -0.29 | 0.50 | — |
| Fatigue | Mid | Post | 8 | 2.3 (0.90-5.4) | 2.3 (0.80-3.8) | -0.40 (-1.3 to 0.40) | -0.44 | 0.31 | — |
| Fatigue | Pre | Post | 8 | 3.1 (2.1-5.3) | 2.3 (0.80-3.8) | -0.70 (-6.9 to 2.9) | -0.21 | 0.69 | — |
| **SWB** | **Pre** | **Mid** | **9** | **26 (23-30)** | **33 (30-38)** | **7.5 (5.5 to 10)** | **1.0** | **0.0039** | **0.012** |
| **SWB** | **Mid** | **Post** | **8** | **36 (30-39)** | **32 (26-35)** | **-3.0 (-9.0 to 0.0)** | **-0.89** | **0.047** | **0.047** |
| SWB | Pre | Post | 8 | 27 (23-30) | 32 (26-35) | 5.2 (-2.0 to 8.0) | 0.61 | 0.15 | 0.15 |
| **PTG** | **Pre** | **Mid** | **9** | **2.8 (1.9-3.6)** | **3.2 (2.7-4.1)** | **1.0 (0.50 to 1.6)** | **1.0** | **0.016** | **0.047** |
| PTG | Mid | Post | 8 | 3.4 (2.8-4.2) | 3.5 (2.8-4.1) | 0.30 (-0.60 to 0.70) | 0.39 | 0.38 | 0.38 |
| **PTG** | **Pre** | **Post** | **8** | **3.0 (2.1-3.6)** | **3.5 (2.8-4.1)** | **1.1 (0.70 to 1.6)** | **1.0** | **0.031** | **0.047** |
