## Supplementary material for "Feasibility and Psychological Outcomes of a Clinical Meditation Intervention for Patients with Cancer: A Mixed-Methods Pilot Study": Multimedia Appendix 3: SRQR Checklist

### Multimedia Appendix 3. SRQR (Standards for Reporting Qualitative Research) Checklist

This checklist documents adherence to the 21-item SRQR guideline for the qualitative component of this mixed-methods pilot study (thematic analysis of free-text questionnaire responses).

*Reference: O'Brien BC, Harris IB, Beckman TJ, Reed DA, Cook DA. Standards for reporting qualitative research: a synthesis of recommendations. Acad Med. 2014;89(9):1245-1251. doi:10.1097/ACM.0000000000000388.*

| # | **Topic** | **Item** | Status | **Location in Manuscript** |
| --- | --- | --- | --- | --- |
| 1 | Title | Concise description of the nature and topic of the study identifying the study as qualitative or indicating the approach (eg, ethnography, grounded theory) or data collection methods (eg, interview, focus group) is recommended | Yes | Title page |
| 2 | Abstract | Summary of key elements of the study using the abstract format of the intended publication; typically includes background, purpose, methods, results, and conclusions | Yes | Abstract (Background, Objective, Methods, Results, Conclusions) |
| 3 | Problem formulation | Description and significance of the problem/phenomenon studied; review of relevant theory and empirical work; problem statement | Yes | Introduction (clinical context, prior research, and study rationale) |
| 4 | Purpose or research question | Purpose of the study and specific objectives or questions | Yes | Introduction, final paragraph (primary feasibility objective and secondary exploratory objectives) |
| 5 | Qualitative approach and research paradigm | Qualitative approach (eg, ethnography, grounded theory, case study, phenomenology, narrative research) and guiding theory if appropriate; identifying the research paradigm is also recommended; rationale | Yes | Introduction, final two paragraphs; Methods, Qualitative Analysis and Mixed-Methods Integration (convergent design and Braun and Clarke thematic analysis). A research paradigm is not explicitly named. |
| 6 | Researcher characteristics and reflexivity | Researchers' characteristics that may influence the research, including personal attributes, qualifications/experience, relationship with participants, assumptions, and/or presuppositions; potential or actual interaction between researchers' characteristics and the research questions, approach, methods, results, and/or transferability | Yes | Methods, Intervention Program and Qualitative Analysis and Mixed-Methods Integration; Discussion, Limitations; Authors’ Contributions. AN’s participant contact and analytic role, NI’s qualitative research expertise, and both AN’s and NI’s completion of Clinical Meditation instructor training are described. AN’s involvement in recruitment and group sessions and its potential influence on responses and interpretation are acknowledged. |
| 7 | Context | Setting/site and salient contextual factors; rationale | Yes | Methods, Participants and Recruitment (Institute of Medical Science, The University of Tokyo Hospital) |
| 8 | Sampling strategy | How and why research participants, documents, or events were selected; criteria for deciding when no further sampling was necessary (eg, sampling saturation); rationale | Partial | Methods, Participants and Recruitment (eligibility, recruitment procedures, target of 10 participants, justified as a pilot study without a formal power calculation); Results, Participant Characteristics and Feasibility and Qualitative Findings (qualitative analyses included all participants who provided free-text responses); Discussion, Limitations. The rationale for the specific target size and the recruitment stopping criterion are not stated, and saturation was not confirmed. |
| 9 | Ethical issues pertaining to human subjects | Documentation of approval by an appropriate ethics review board and participant consent, or explanation for lack thereof; other confidentiality and data security issues | Yes | Methods, Ethical Considerations; Qualitative Analysis and Mixed-Methods Integration (removal of personally identifying information before LLM input and settings excluding the data from model training). |
| 10 | Data collection methods | Types of data collected; details of data collection procedures including (as appropriate) start and stop dates of data collection and analysis, iterative process, triangulation of sources/methods, and modification of procedures in response to evolving study findings; rationale | Yes | Methods, Questionnaire Data Collection and Qualitative Analysis and Mixed-Methods Integration (web-based free-text responses; Q1 at Pre/Mid/Post and Q2 at Mid/Post; individual-level integration). Calendar dates of collection are not stated. |
| 11 | Data collection instruments and technologies | Description of instruments (eg, interview guides, questionnaires) and devices (eg, audio recorders) used for data collection; if/how the instrument(s) changed over the course of the study | Yes | Methods, Questionnaire Data Collection and Qualitative Analysis and Mixed-Methods Integration (web-based questionnaire; verbatim Q1/Q2 wording and administration time points) |
| 12 | Units of study | Number and relevant characteristics of participants, documents, or events included in the study; level of participation (could be reported in results) | Yes | Results, Participant Characteristics and Feasibility and Qualitative Findings (8 participants, 34 free-text responses, response lengths, and 72 coded passages; Patient_H provided no free-text responses) |
| 13 | Data processing | Methods for processing data prior to and during analysis, including transcription, data entry, data management and security, verification of data integrity, data coding, and anonymization/deidentification of excerpts | Yes | Methods, Ethical Considerations and Qualitative Analysis and Mixed-Methods Integration (coded identifiers, separately stored key, removal of personally identifying information before LLM input, settings excluding these data from model training, analysis of Japanese-language responses, coding, and AN’s verification against all original responses). Quotations were translated from Japanese into English using an LLM, and the translations were checked by AN; see also Multimedia Appendix 4. Audio transcription is not applicable to written responses. |
| 14 | Data analysis | Process by which inferences, themes, etc, were identified and developed, including the researchers involved in data analysis; usually references a specific paradigm or approach; rationale | Yes | Methods, Qualitative Analysis and Mixed-Methods Integration (LLM-assisted candidate coding, codebook, recoding and theme development; AN’s source checks and revisions; NI’s review; participant-level joint display) |
| 15 | Techniques to enhance trustworthiness | Techniques to enhance trustworthiness and credibility of data analysis (eg, member checking, audit trail, triangulation); rationale | Yes | Methods, Qualitative Analysis and Mixed-Methods Integration; Authors’ Contributions. AN checked codes, themes, and quotations against source material; NI reviewed the codebook and themes and advised on refinements. This was not independent re-coding or a full audit. |
| 16 | Synthesis and interpretation | Main findings (eg, interpretations, inferences, and themes); might include development of a theory or model, or integration with prior research or theory | Yes | Results, Qualitative Findings and Integrating Quantitative and Qualitative Findings at the Individual Level; Figure 4; Discussion, Principal Findings |
| 17 | Links to empirical data | Evidence (eg, quotes, field notes, text excerpts, photographs) to substantiate analytic findings | Yes | Results, Qualitative Findings (direct participant quotations for each theme) |
| 18 | Integration with prior work, implications, transferability, and contribution(s) to the field | Short summary of main findings; explanation of how findings and conclusions connect to, support, elaborate on, or challenge conclusions of earlier scholarship; discussion of scope of application/generalizability; identification of unique contribution(s) to scholarship in a discipline or field | Yes | Discussion, Comparison With Prior Work and Limitations (theoretical interpretation, prior studies, contribution, and limits to transferability) |
| 19 | Limitations | Trustworthiness and limitations of findings | Yes | Discussion, Limitations |
| 20 | Conflicts of interest | Potential sources of influence or perceived influence on study conduct and conclusions; how these were managed | Yes | Conflicts of Interest; Methods, Intervention Program and Qualitative Analysis and Mixed-Methods Integration; Discussion, Limitations (developer/instructor roles, noninvolvement of DO/TK in interpretation, AN’s dual role and potential bias, and NI’s review) |
| 21 | Funding | Sources of funding and other support; role of funders in data collection, interpretation, and reporting | Yes | Funding (Operating Expense Grants from the University of Tokyo and the Future Medicine Development Fund of the Institute of Medical Science, the University of Tokyo; the latter is supported by private donations). The funders had no role in data collection, interpretation of the findings, or preparation of the manuscript. |

*Note: "Partial" indicates that additional reporting or author clarification is needed for sampling rationale and recruitment stopping criteria (Item 8). These ratings concern reporting completeness, not a numerical assessment of study quality.*
